# STREAM Sequential Transformer for Risk Estimation from Asynchronous Medical Events for Dynamic Postoperative AKI Prediction

**DOI:** 10.64898/2026.11.21.26368665

**Authors:** Shuyu Lu, Xiaoyang Ruan, Liwei Wang, Jaerong Ahn, Yafen Liang, Xiaomeng Wang, Andrew Wen, Jun Jiang, Yian Hu, Xin Li, Hongfang Liu

**Affiliations:** McWilliams School of Biomedical Informatics, The University of Texas Health Science Center at Houston, Houston, TX, USA; Department of Quantitative and Systems Health Sciences, Dell Medical School, The University of Texas at Austin, Austin, TX, USA; Department of Anesthesiology, Critical Care and Pain Medicine, McGovern Medical School, The University of Texas Health Science Center at Houston, Houston, TX, USA

## Abstract

Postoperative acute kidney injury (PO-AKI) risk estimates evolve as clinical information accumulates, yet many electronic health record (EHR) models use static or fixed-window representations. We developed STREAM, a perioperative event-stream Transformer that represents irregular EHR data as chronological clinical events and supports surgery-index prediction at surgery completion and event-by-event postoperative AKI prediction. In a retrospective cohort of 167,078 noncardiac surgical episodes, STREAM achieved surgery-index AUROC/AUPRC of 0.942/0.771, the Time-series Transformer 0.933/0.729, and GRU-D 0.926/0.695. Postoperative macro-AUPRC across 6-hour evaluation intervals was 0.589, 0.525, and 0.547, respectively. STREAM showed more consistent calibration across postoperative time points than the comparators. Event-arrival attribution and within-window permutation analysis enabled event-level interpretation of how newly arrived information and fine-grained event ordering shaped evolving risk estimates. These findings support event-stream modeling as an interpretable framework for dynamic, temporally resolved prediction from longitudinal EHR data as well as a potential risk-estimation component for future perioperative digital twins.

## Introduction

Acute kidney injury (AKI) is a common and clinically important complication following major surgeries. Depending on the surgical population and clinical setting, the incidence of postoperative AKI (PO-AKI) has been reported to range from approximately 3% to 40% ^1–3^. PO-AKI has been regarded as a sentinel surgical complication because it is associated with increased short-term morbidity and mortality as well as longer-term adverse outcomes, including chronic kidney disease, intensive care utilization, and hospital readmission ^4,5^. PO-AKI risk reflects the interaction of baseline patient characteristics, surgical factors, and evolving perioperative conditions such as hemodynamic instability, volume status, medication exposure, and postoperative complications ^6^.

Although renal injury is particularly well recognized after cardiac surgery, AKI following noncardiac surgery affects a broader and heterogeneous surgical population and may be less consistently recognized ^7,8^. As noncardiac surgical patients are not always subject to systematic renal surveillance, PO-AKI in this setting may be more likely to be under-recognized. This creates a need for accurate and timely risk stratification across broader non-cardiac surgical populations, especially when AKI risk is not apparent at baseline. This dynamic and multifactorial risk profile makes PO-AKI a useful setting for evaluating models that can update risk as perioperative information accumulates.

Previous PO-AKI prediction studies have used clinical risk scores, conventional machine-learning models, and more recently deep-learning approaches to identify high-risk patients ^9–11^. Many of these approaches rely on static or aggregated perioperative variables and primarily estimate risk at a single time point. Sequential models such as recurrent neural networks and GRU-D extend prediction across longitudinal EHR data by incorporating temporal patterns and missingness ^12,13^. However, structured EHR observations are inherently irregular: clinical events occur at unequal intervals, multiple events may occur within short periods, and information density can change substantially over the course of a clinical trajectory. Fixed-window representations simplify this structure by aggregating observations into predefined intervals and often applying imputation, potentially removing information about fine-grained timing, repeated events, and within-window ordering.

Event-based and token-based representations provide an alternative approach to longitudinal EHR modeling. STraTS represents sparse and irregular clinical observations using time– variable–value representations ^14^, while Event Stream GPT models complex clinical trajectories as continuous-time sequences of events ^15^. At a larger scale, Curiosity models longitudinal EHRs as sequences of medical events and generates plausible future event trajectories to support risk estimation across diverse clinical tasks ^16^. Together with other event-based approaches, this work demonstrates the predictive potential of preserving clinical histories as event sequences. Related models such as BEHRT, Med-BERT, and CEHR-BERT have shown that structured clinical histories can be represented as sequences of clinical concepts with contextual and temporal information ^17–19^. Recent benchmarking has further shown competitive performance of event-stream representations across multiple clinical datasets^20^. The remaining question is therefore not whether EHR data can be represented as event sequences, but what is gained by retaining event-level temporal structure and how that information contributes to dynamic prediction. In particular, the effects of within-window event ordering, event-by-event updating, and longitudinal clinical context remain incompletely characterized ^21^.

The perioperative setting provides a useful test bed for these questions because the surgery forms a clear temporal anchor between the preoperative state and an asynchronously evolving postoperative course. We therefore developed STREAM (<u>S</u>equential <u>T</u>ransformer for <u>R</u>isk <u>E</u>stimation from <u>A</u>synchronous <u>M</u>edical Events) for dynamic prediction of coded PO-AKI following noncardiac surgery. STREAM represents each surgical episode (an eligible index surgery and its associated perioperative EHR event stream) as a chronological sequence of heterogeneous clinical events, uses a dedicated surgery-index representation to summarize information available at surgery, and updates postoperative risk as new events enter the stream.

Our contributions are as follows. First, we developed STREAM, a perioperative event-stream Transformer that represents heterogeneous EHR data as a chronological sequence of clinical events and supports both surgery-index (i.e. right after surgery) risk estimation and event-by-event postoperative updating to reassess PO-AKI risk as new clinical information becomes available. Second, we systematically compared event-stream modeling with static and fixed-window time-series representations for coded PO-AKI prediction, evaluating surgery-index discrimination, postoperative dynamic discrimination, and calibration across postoperative time. Third, we investigated how event-level temporal structure contributed to model behavior through architectural and input ablations, event-arrival attribution, within-window order perturbation, temporal grouped deletion, and age and surgery substitution. Together, these analyses connect predictive performance of STREAM with the longitudinal clinical context and event ordering underlying dynamic risk updates.

## Results

### Study Cohort Characteristics

The analytic cohort comprised 167,078 adult noncardiac surgical episodes from 134,741 patients across six surgical categories (**Figure 1A**). Each episode represented an eligible surgery and its associated perioperative EHR events. The chronological split yielded 143,540 episodes in the development set and 23,538 in the held-out temporal test set, of which 8,147 (5.7%) and 2,371 (10.1%), respectively, had a recorded PO-AKI diagnosis within 30 days after surgery.

**Figure 1.**
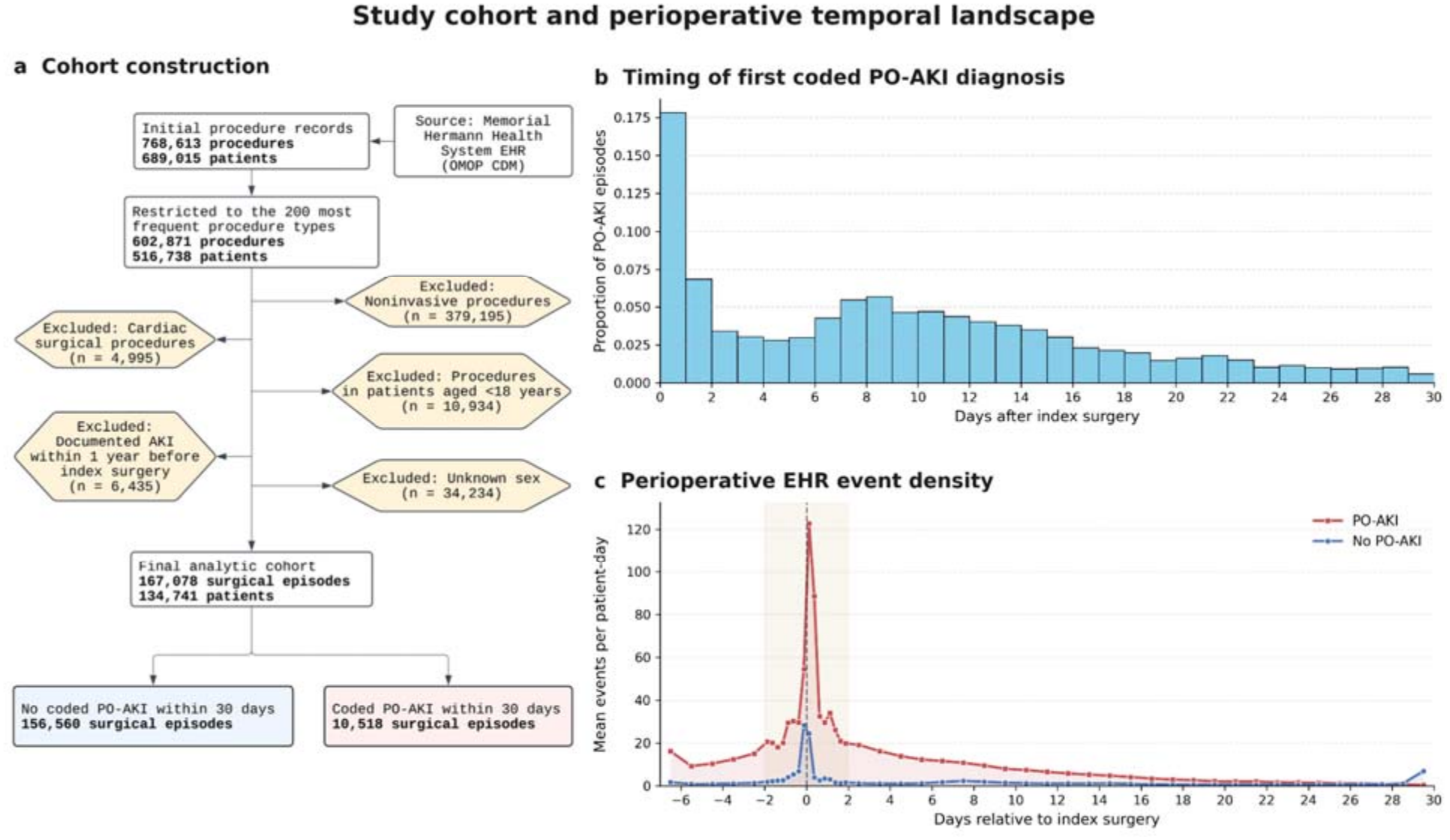
Study cohort and perioperative temporal landscape. **(a)** Cohort construction. Surgical episodes were identified from the Memorial Hermann Health System EHR mapped to the OMOP Common Data Model and sequentially filtered by procedure frequency, invasiveness, cardiac surgery status, age, prior documented AKI, and sex availability. The final analytic cohort comprised 167,078 surgical episodes from 134,741 patients, including 10,518 episodes with coded PO-AKI within 30 days after surgery. **(b)** Distribution of the first coded PO-AKI diagnosis by postoperative day among PO-AKI episodes. **(c)** Mean perioperative EHR event density in episodes with and without coded PO-AKI, aligned to the index surgery. Both coded PO-AKI diagnoses and EHR event activity were concentrated in the early postoperative period.

Table 1 summarizes cohort characteristics. Across both sets, patients in episodes with coded PO-AKI were older and more frequently male, and abdominal surgery was the most common surgical category. Skin surgery, limited here to split-thickness skin grafting and skin-flap revision, was proportionally more common in episodes with coded PO-AKI. Episodes with coded PO-AKI also contained more condition and medication events than those without coded PO-AKI, whereas laboratory events were relatively sparse in both groups and less frequent in episodes with coded PO-AKI.

**Table 1.**
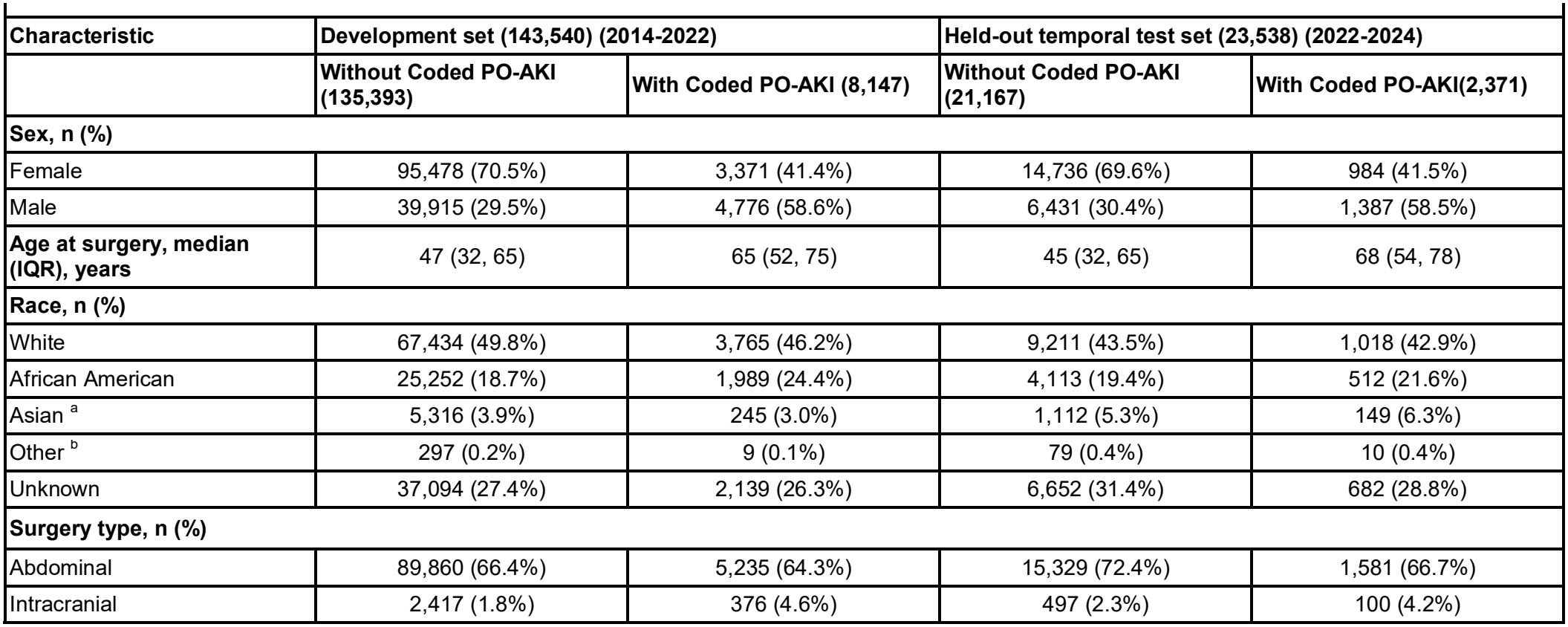
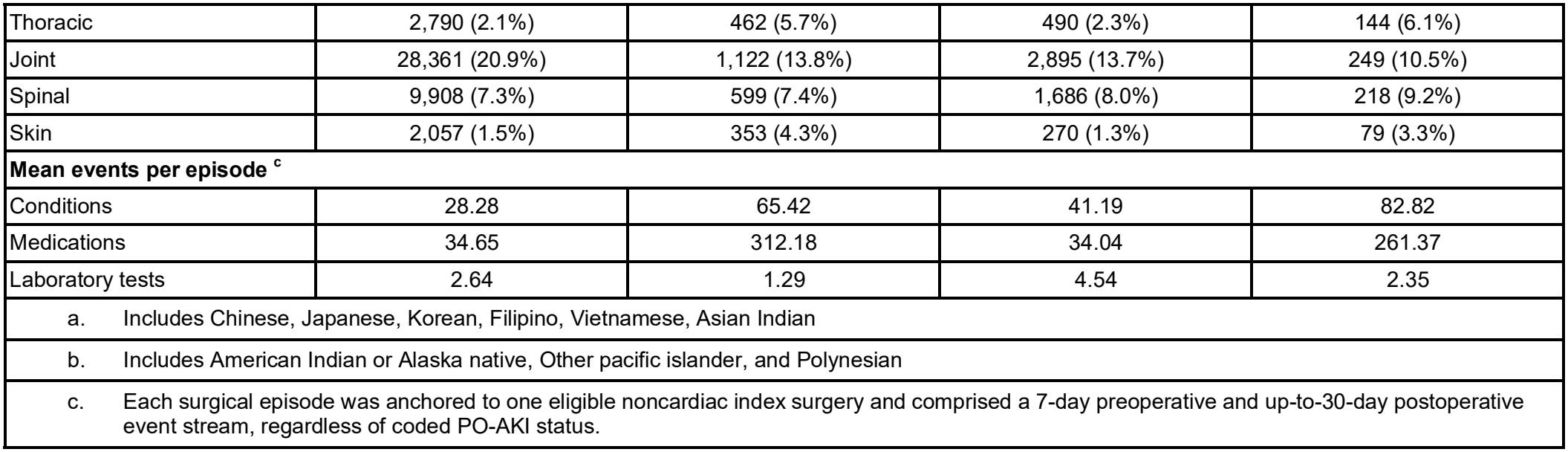
Study Cohort Characteristics.

| **Characteristic** | **Development set (143,540) (2014-2022)** |  | **Held-out temporal test set (23,538) (2022-2024)** |  |
|---|---|---|---|---|
|  | **Without Coded PO-AKI (135,393)** | **With Coded PO-AKI (8,147)** | **Without Coded PO-AKI (21,167)** | **With Coded PO-AKI(2,371)** |
| **Sex, n (%)** |  |  |  |  |
| Female | 95,478 (70.5%) | 3,371 (41.4%) | 14,736 (69.6%) | 984 (41.5%) |
| Male | 39,915 (29.5%) | 4,776 (58.6%) | 6,431 (30.4%) | 1,387 (58.5%) |
| **Age at surgery, median (IQR), years** | 47 (32, 65) | 65 (52, 75) | 45 (32, 65) | 68 (54, 78) |
| **Race, n (%)** |  |  |  |  |
| White | 67,434 (49.8%) | 3,765 (46.2%) | 9,211 (43.5%) | 1,018 (42.9%) |
| African American | 25,252 (18.7%) | 1,989 (24.4%) | 4,113 (19.4%) | 512 (21.6%) |
| Asian<sup>a</sup> | 5,316 (3.9%) | 245 (3.0%) | 1,112 (5.3%) | 149 (6.3%) |
| Other<sup>b</sup> | 297 (0.2%) | 9 (0.1%) | 79 (0.4%) | 10 (0.4%) |
| Unknown | 37,094 (27.4%) | 2,139 (26.3%) | 6,652 (31.4%) | 682 (28.8%) |
| **Surgery type, n (%)** |  |  |  |  |
| Abdominal | 89,860 (66.4%) | 5,235 (64.3%) | 15,329 (72.4%) | 1,581 (66.7%) |
| Intracranial | 2,417 (1.8%) | 376 (4.6%) | 497 (2.3%) | 100 (4.2%) |
| Thoracic | 2,790 (2.1%) | 462 (5.7%) | 490 (2.3%) | 144 (6.1%) |
| Joint | 28,361 (20.9%) | 1,122 (13.8%) | 2,895 (13.7%) | 249 (10.5%) |
| Spinal | 9,908 (7.3%) | 599 (7.4%) | 1,686 (8.0%) | 218 (9.2%) |
| Skin | 2,057 (1.5%) | 353 (4.3%) | 270 (1.3%) | 79 (3.3%) |
| **Mean events per episode<sup>c</sup>** |  |  |  |  |
| Conditions | 28.28 | 65.42 | 41.19 | 82.82 |
| Medications | 34.65 | 312.18 | 34.04 | 261.37 |
| Laboratory tests | 2.64 | 1.29 | 4.54 | 2.35 |

a. Includes Chinese, Japanese, Korean, Filipino, Vietnamese, Asian Indian
b. Includes American Indian or Alaska native, Other pacific islander, and Polynesian
c. Each surgical episode was anchored to one eligible noncardiac index surgery and comprised a 7-day preoperative and up-to-30-day postoperative event stream, regardless of coded PO-AKI status.

### Temporal Distribution of Coded PO-AKI and Perioperative EHR Events

As shown in **Figure 1b**, first coded PO-AKI diagnoses were concentrated early after surgery, with the largest proportion recorded on the day of surgery and a smaller secondary peak approximately 1 week later. Perioperative EHR activity was similarly concentrated around the index surgery (**Figure 1c**), with mean event density peaking near *t*_0_ =0 and declining thereafter.

Episodes with coded PO-AKI showed higher event density than episodes without coded PO-AKI, particularly around the time of surgery. Overall, both coded outcome documentation and EHR event density were concentrated in the early postoperative period, with substantially sparser observations at later time points.

### Predictive Performance at the Surgery-Index and During Postoperative Monitoring

STREAM generated the surgery-index prediction from a dedicated learnable [*BASE*] token that summarized information available at the index surgery; postoperative event representations then produced updated risk estimates as new events were recorded. At the surgery-index, STREAM achieved the highest discrimination among the evaluated models, with a mean AUROC of 0.942 and AUPRC of 0.771 (**Table 2**). The strongest fixed-window comparator, the Time-series Transformer, achieved an AUROC of 0.933 and AUPRC of 0.729, whereas XGBoost achieved an AUROC of 0.907 and AUPRC of 0.605.

**Table 2.** Surgery-index and postoperative dynamic predictive performance.

| **Model Type** | **Model** | **Surgery-index AUROC** | **Surgery-index AUPRC** | **Dynamic macro-AUROC** | **Dynamic macro-AUPRC** |
|---|---|---|---|---|---|
| Static | Logistic Regression | 91.05 (91.02, 91.07) | 61.17 (61.08, 61.26) | — | — |
|  | XGBoost | 90.69 (90.57, 90.88) | 60.47 (60.24, 60.68) | — | — |
| Fixed-window Time Series | LSTM | 92.42 (92.21, 92.56) | 67.76 (66.61, 68.34) | 93.24 (92.79, 93.59) | 44.49 (41.97, 46.35) |
|  | GRU-D | 92.63 (92.51, 92.83) | 69.48 (69.04, 70.28) | 94.23 (93.98, 94.39) | 54.70 (53.37, 55.41) |
|  | Time-series Transformer | 93.33 (93.27, 93.41) | 72.88 (72.72, 73.07) | 94.76 (94.59, 94.86) | 52.45 (50.72, 53.85) |
| Event Stream | STREAM | 94.19 (94.02, 94.32) | 77.14 (76.70, 77.73) | 94.82 (94.72, 94.96) | 58.86 (58.06, 60.23) |

Values are mean (minimum, maximum) across five fold-specific models evaluated on the same held-out temporal test set.

STREAM also achieved the highest postoperative macro-averaged discrimination across valid 6-hour intervals over the 30-day follow-up. Its macro-AUROC was 0.948 and macro-AUPRC was 0.589, compared with 0.948 and 0.525 for the Time-series Transformer and 0.942 and 0.547 for GRU-D (**Table 2**). Because macro metrics were calculated within each 6-hour interval and then averaged equally across valid intervals, these results summarize within-time-point discrimination across the postoperative trajectory. Complete surgery-index and postoperative performance metrics are provided in **Supplementary Tables S1 and S2**, respectively.

AUPRC evaluated at successive 6-hour postoperative intervals showed distinct performance patterns during early and later postoperative follow-up (**Figure 2**). At each interval, the risk set comprised episodes still free of coded PO-AKI; an episode in this set was labeled positive if it subsequently received a qualifying diagnosis by postoperative day 30. During the first 48 hours after surgery, STREAM consistently achieved the highest AUPRC, remaining approximately 0.75–0.79 compared with approximately 0.72–0.75 for the Time-series Transformer, with clearer separation from the fixed-window models during this event-dense period. After 48 hours, AUPRC declined across all models as the number of remaining coded PO-AKI-positive episodes and the density of newly recorded EHR events decreased. STREAM generally maintained higher AUPRC across the later postoperative period, although differences between models became less consistent and estimates more variable as the number of positive episodes in the remaining risk set decreased toward postoperative day 30.

**Figure 2.**
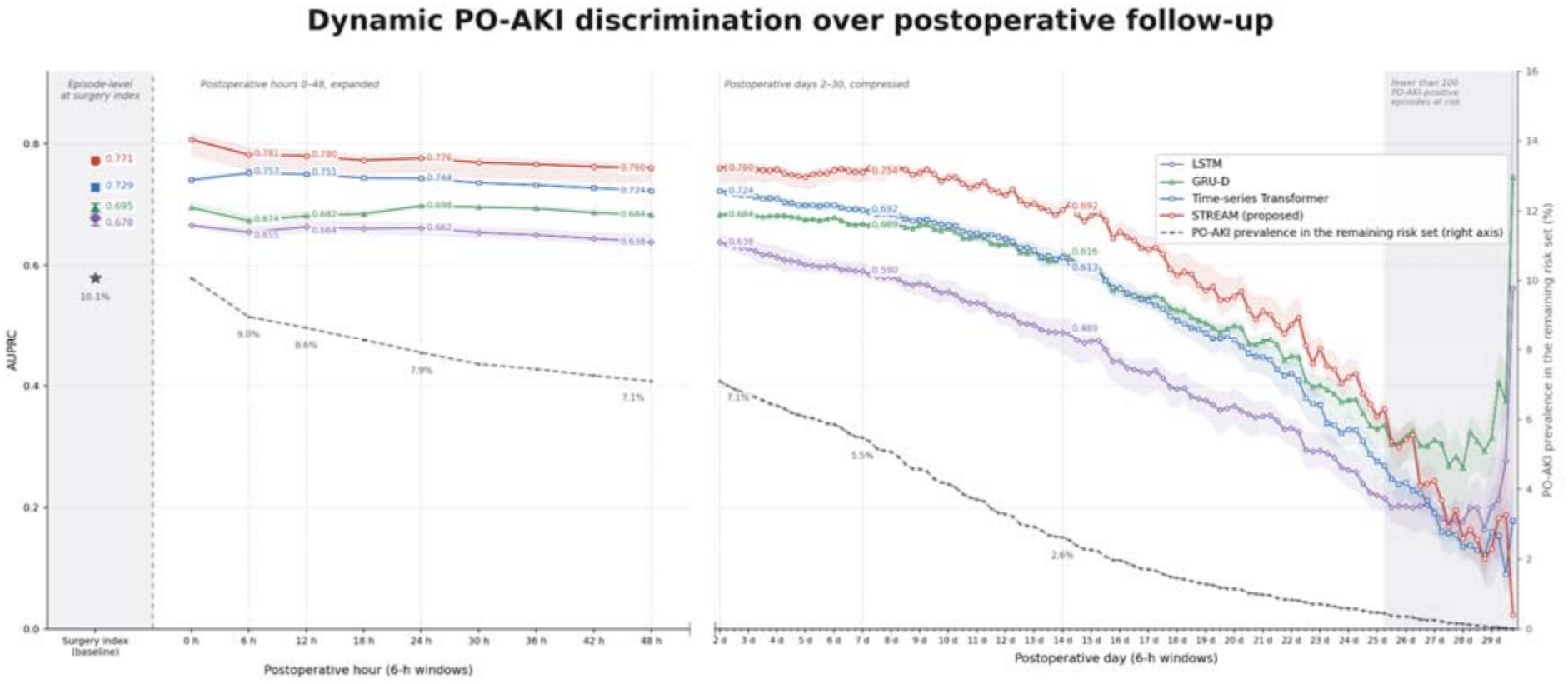
Temporal evolution of PO-AKI discrimination from the surgery-index through 30-day postoperative monitoring. AUPRC was evaluated at the surgery-index and across sequential 6-hour postoperative windows for STREAM and the fixed-window time-series models. The first 48 postoperative hours are shown at expanded 6-hour resolution, whereas days 2–30 are displayed on a compressed day scale while retaining the 6-hour evaluation windows. Solid lines show the mean across five fold-specific models, with shaded bands indicating the fold-wise minimum–maximum range. The dashed line on the secondary axis shows coded PO-AKI prevalence in the remaining risk set over time. Late intervals with fewer than 100 coded PO-AKI-positive episodes remaining at risk are shaded separately.

### Calibration Performance

STREAM achieved lower Brier scores across the evaluated time points and showed less pronounced early postoperative overprediction than the fixed-window models, although residual miscalibration remained (**Figure 3**). At the surgery-index, its predicted probabilities closely followed the 45° identity line over the range containing most episodes. Calibration remained relatively stable at 6, 12, 24, and 48 hours and postoperative day 7, with larger deviations occurring in the sparsely populated high-risk range. In comparison, the fixed-window time-series models showed greater overprediction during the early postoperative period, particularly at 6 and 12 hours, with closer agreement at later time points. Across models, predicted-risk distributions became increasingly concentrated in the low-risk range over postoperative follow-up. Quantitative calibration measures, including Brier score, Brier skill score, calibration intercept, and calibration slope, are reported in **Supplementary Table S3**.

**Figure 3.**
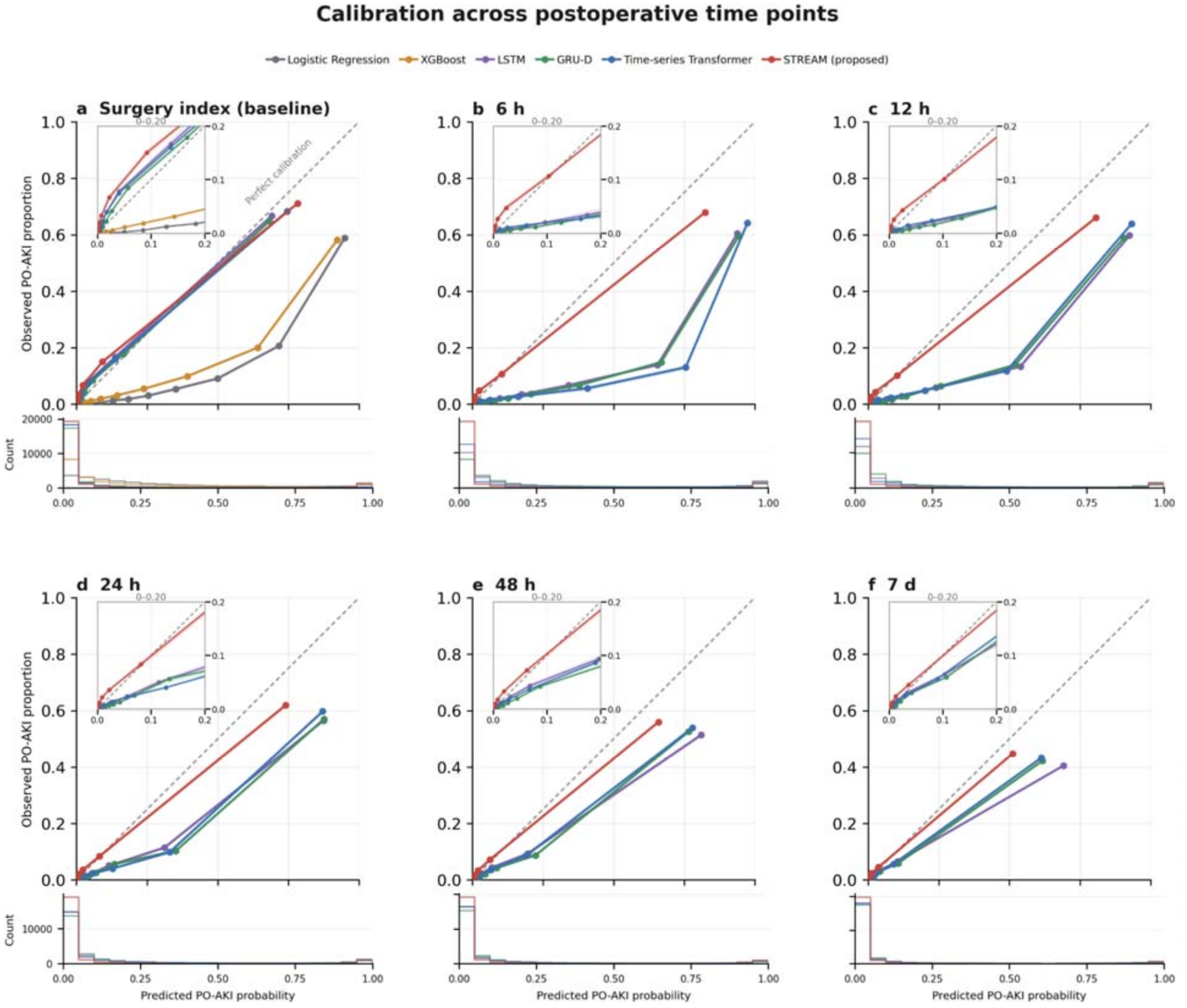
Calibration of PO-AKI risk prediction across postoperative time points. Calibration curves compare mean predicted PO-AKI probability with the observed coded PO-AKI proportion at the surgery-index and at 6, 12, 24, 48 hours, and 7 days after surgery. Solid lines represent the mean across five fold-specific models evaluated on the same held-out temporal test set. Insets enlarge the low-risk range (predicted probability, 0–0.20), and histograms below each panel show the corresponding distributions of predicted probabilities. The dashed 45° identity line represents perfect calibration. Static models were evaluated only at the surgery-index baseline.

### Ablation Study Results

Ablation analysis showed that the dedicated *[BASE]* representation had the largest architecture-level effect on postoperative discrimination (**Table 3**). Replacing *[BASE]* with the surgery token reduced dynamic macro-AUPRC from 0.589 to 0.560, whereas replacing structured block attention with unidirectional attention produced a smaller decrease to 0.583. Removing continuous time encoding did not reduce postoperative macro-AUPRC in this evaluation, whereas removal of the masked event modeling (MEM) objective, which reconstructs selected preoperative event codes during training, produced only a small decrease. Removing the surgery-index prediction loss substantially reduced surgery-index AUPRC from 0.771 to 0.154 and also modestly reduced postoperative macro-AUPRC to 0.564.

**Table 3.** Ablation analysis of STREAM.

| Category | Ablation | Surgery-index AUPRC | Dynamic macro-AUPRC |
|---|---|---|---|
| Full model | — | 77.14 | 58.86 |
| Architecture | No time encoding (A1) | 76.37 (−0.77) | 59.27 (+0.41) |
|  | Unidirectional attention (A2) | 77.33 (+0.19) | 58.31 (−0.55) |
|  | Surgery token as baseline (A3) | 76.91 (−0.24) | 55.95 (−2.91)* |
| Training objective | No baseline loss (A4) | 15.36 (−61.78)*† | 56.39 (−2.47) |
|  | No MEM (Masked Event Modeling) loss (A5) | 76.83 (−0.31) | 58.20 (−0.66) |
| Input modality | No medication events (I1) | 52.49 (−24.65)* | 23.10 (−35.76)* |
|  | No laboratory events (I2) | 78.31 (+1.17) | 56.77 (−2.09) |
|  | No condition events (I3) | 74.25 (−2.89)* | 46.61 (−12.25)* |
|  | Generic surgery token (I4) | 77.29 (+0.15) | 58.20 (−0.66) |
| Interaction | Surgery-as-baseline + generic surgery (A3+I4) | 76.60 (−0.54) | 57.47 (−1.39) |

† The no-baseline-loss model did not train the surgery-index prediction head; its surgery-index AUPRC is therefore shown only to demonstrate dependence of the baseline head on explicit supervision.
\* Lower than the full model in all five folds. Values are five-fold means (%); values in parentheses are the mean paired within-fold change from the full model in percentage points, computed before rounding. Dynamic macro-AUPRC was calculated separately within each valid 6-hour postoperative interval and summarized as the unweighted mean across intervals.

Among input ablations, removing medication events produced the largest performance decline, reducing surgery-index AUPRC from 0.771 to 0.525 and postoperative macro-AUPRC from 0.589 to 0.231. Removing condition events also reduced both surgery-index and postoperative performance, with macro-AUPRC decreasing to 0.466. Removal of laboratory events and replacement of surgery identity with a generic surgery token produced smaller changes. In the joint A3+I4 ablation, macro-AUPRC was 0.575, partially attenuating the decrease observed when the surgery token alone replaced *[BASE]*.

### Interpretability Evaluation

Interpretability analyses addressed three complementary questions: which clinical information contributed to model predictions, how risk estimates changed as new postoperative events arrived, and whether fine-grained event ordering affected prediction. The primary analyses focused on event-arrival attribution and within-window order sensitivity, with perturbation-based analyses of age, surgery identity, and temporally grouped clinical events provided in the Supplementary material.

### Event-Arrival Attribution

After accounting for unresolved ordering among events sharing the same recorded timestamp, code-level event-arrival attribution identified clinical events associated with increases and decreases in STREAM’s postoperative PO-AKI risk estimates (**Figure 4**). Codes associated with increases included renal, metabolic, cardiovascular, and perioperative medication-related codes, whereas those associated with decreases more often involved obstetric and gynecologic conditions. These estimates represent the average direction and magnitude of event-associated changes in model logit during the first 7 postoperative days.

**Figure 4.**
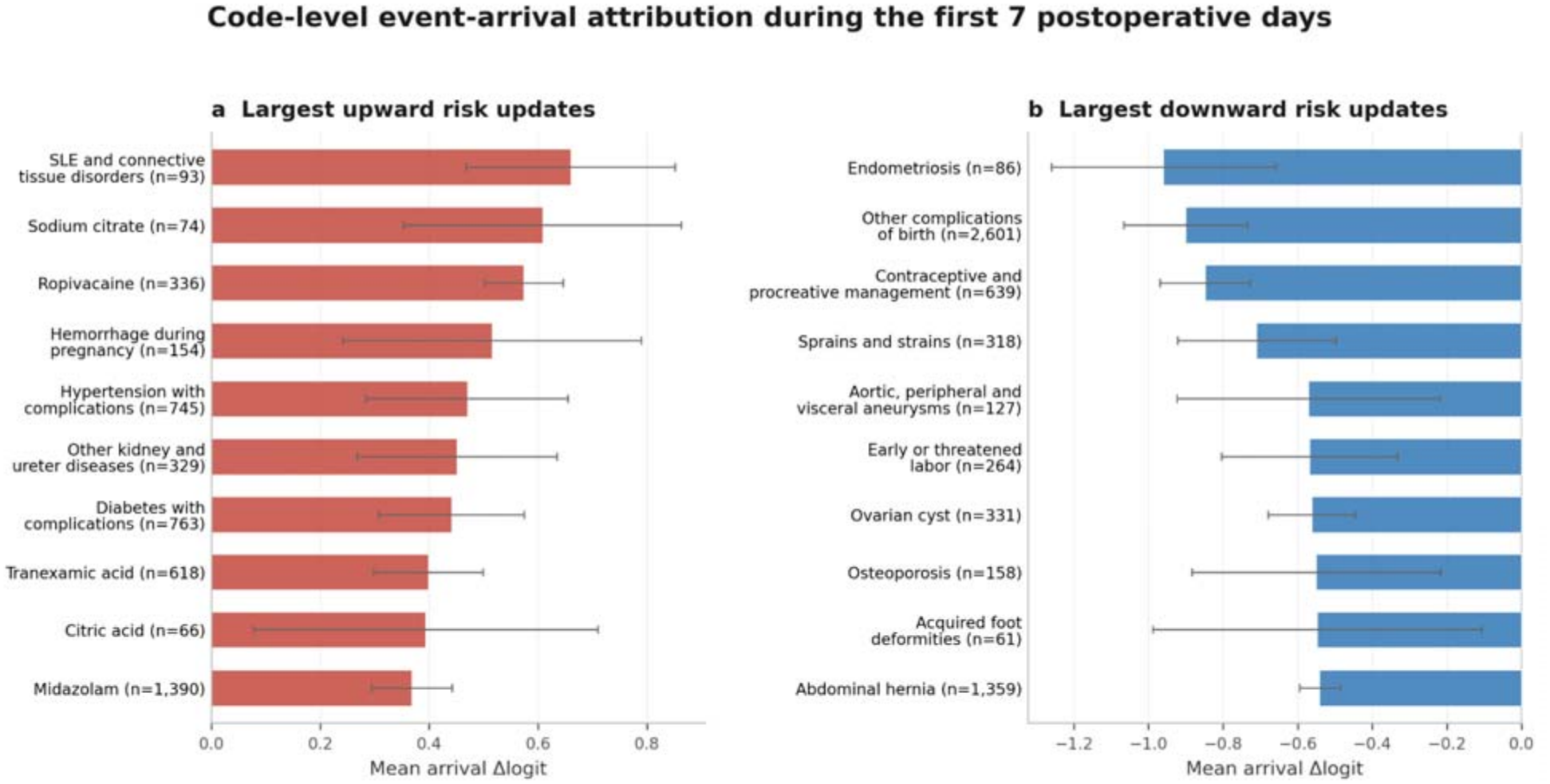
Code-level event-arrival attribution during the first 7 postoperative days. Clinical codes were ranked by their mean event-arrival attribution across five fold-specific STREAM models. **(a)** Codes associated with the largest increases in model-estimated PO-AKI risk after event arrival. **(b)** Codes associated with the largest decreases in model-estimated PO-AKI risk after event arrival. Attribution was quantified as the change in PO-AKI logit associated with event arrival; for events sharing the same recorded timestamp, contributions were averaged across within-timestamp permutations. Bars show the five-fold mean attribution, error bars indicate the fold-wise minimum–maximum range, and labels report the number of contributing patients. Only codes with concordant attribution direction across all five models are shown.

Representative patient-level trajectories further illustrated how event-specific updates accumulated over postoperative time (**Supplementary Figure S1**). In an episode with coded PO-AKI following laparoscopic appendectomy, predicted risk increased from approximately 0.02 to 1.00 before the first coded PO-AKI diagnosis at 15.4 hours. In an episode without coded PO-AKI following exploratory laparotomy, predicted risk decreased from approximately 0.98 toward 0 over the postoperative trajectory. Matched trajectories from the fixed-window Time-series Transformer for the same episodes are shown in **Supplementary Figure S2**.

### Within-Window Order Sensitivity

Within-window permutation analysis showed that STREAM predictions were sensitive to the fine-grained ordering of postoperative events, with sensitivity varying by both event density and time since surgery (**Figure 5a**). During the first 6 postoperative hours, median order sensitivity increased from 0.25 in windows containing 3–4 events to 0.48 in windows containing at least 33 events. Across the four postoperative periods, windows with at least 33 events had median sensitivities of 0.40–0.50, compared with 0.20–0.27 in windows with 3–8 events. Sensitivity generally increased with event density, although the pattern was not monotonic in every time stratum.

**Figure 5.**
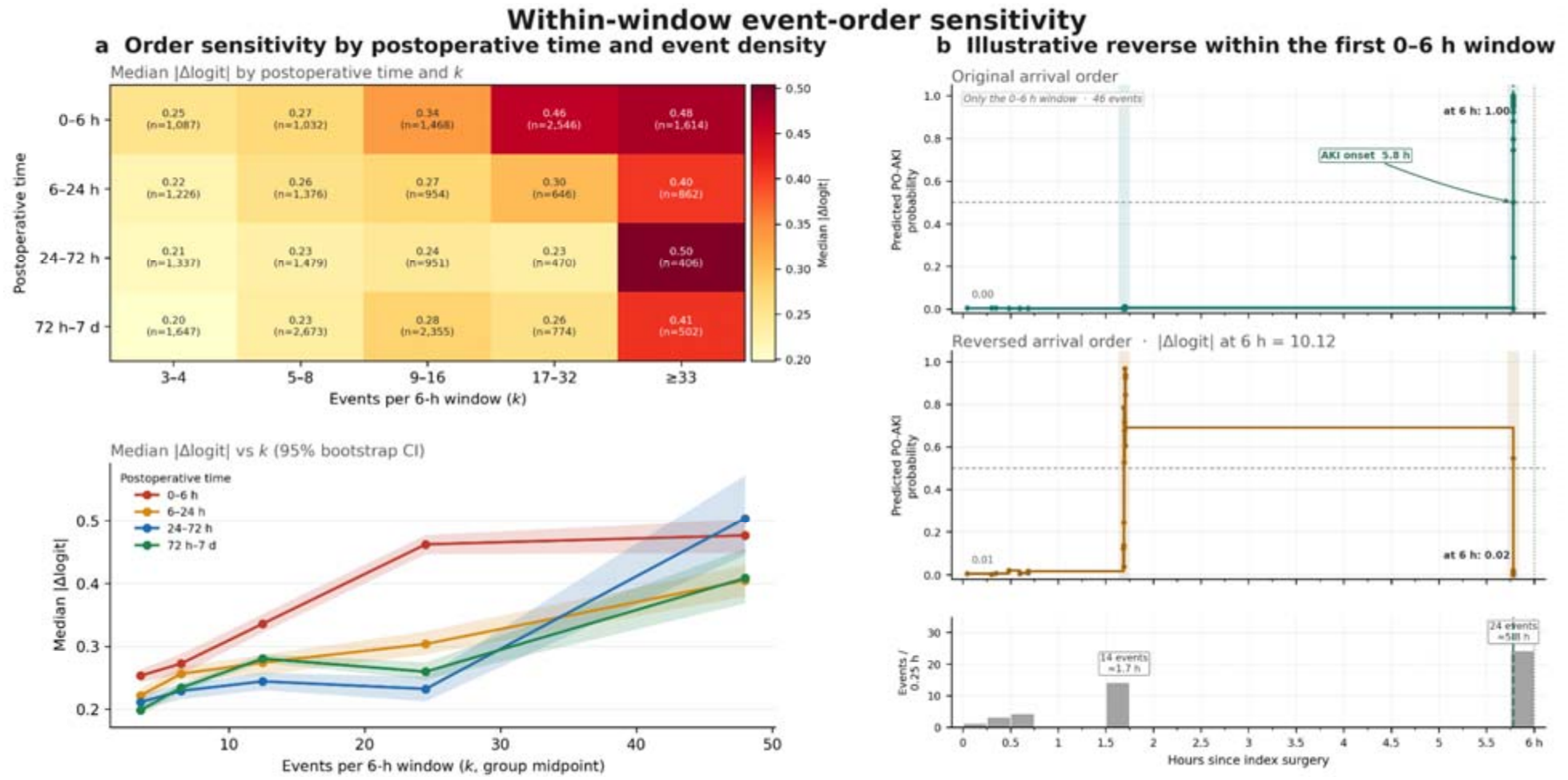
Within-window event-order sensitivity of STREAM. **(a)** Sensitivity across postoperative time and event density. For each eligible 6-hour window, sensitivity was the mean absolute difference in end-of-window PO-AKI logit between the original event order and up to 20 random within-window permutations. Heatmap cells report the median sensitivity and number of contributing windows in each stratum. The line plot shows the same medians with 95% bootstrap confidence intervals from 2,000 window-level resamples. **(b)** Illustrative reversal of event order within a 0–6-hour postoperative window containing 46 events. The same event identities and timestamp positions were retained, but the identities were assigned to those positions in reverse order. End-of-window predicted PO-AKI probabilities were approximately 1.00 in the original order and 0.02 in the reversed order (absolute logit difference, 10.12). The lower plot shows event counts in 0.25-hour bins; the annotated line marks AKI onset at approximately 5.8 hours.

An illustrative full reversal test further demonstrated the potential effect of event ordering on an individual prediction. The same 46 events in one 0–6-hour window were assigned in reverse order to the window’s recorded temporal positions. The predicted PO-AKI probability at the end of the window changed from approximately 1.00 under the original order to 0.02 under the reversed order, an absolute logit difference of 10.12 (**Figure 5b**).

### Supplementary Perturbation-Based Attribution Analyses

Complementary perturbation analyses characterized STREAM’s reliance on age, surgery identity, and temporally grouped clinical events (**Supplementary Figure S3**). Age substitution showed a monotonic increase in surgery-index PO-AKI risk with increasing age. Surgery-substitution effects were positively correlated with observed surgery-specific coded PO-AKI incidence (Spearman *ρ* = 0.667), while temporal grouped deletion identified time-dependent attribution patterns across perioperative periods.

Among codes replicated by both analyses, event-arrival and grouped-deletion attributions were strongly concordant (Spearman *ρ* = 0.822; 91.9% directional agreement), indicating that the two approaches captured related aspects of model behavior while operating at different temporal scales.

## Discussion

In this study, we developed STREAM, a perioperative event-stream Transformer that represents longitudinal EHR data as a chronological sequence of clinical events and updates postoperative risk as new information becomes available. Across a large surgical cohort, STREAM achieved better surgery-index prediction of coded PO-AKI than both static and fixed-window time-series models and better postoperative dynamic discrimination than the fixed-window time-series models. It also showed closer agreement between predicted and observed outcome frequencies across postoperative time points. Event-arrival attribution showed how predictions changed as new clinical information entered the stream, while within-window permutation analysis demonstrated that predictions were sensitive to the fine-grained ordering of events. Together with the ablation analyses, these findings extend prior work on event-based EHR modeling and suggest that event-level representation, longitudinal context, and within-window ordering contribute to dynamic EHR prediction modeling.

A central finding of this study concerns the representation of longitudinal EHR data. Prior work has progressively moved structured EHR modeling from static feature engineering toward sequential and event-based representations. BEHRT and Med-BERT represent coded clinical histories as token sequences, GRU-D explicitly models irregular observation and missingness, and CEHR-BERT incorporates temporal information into structured EHR representations ^17–19,22^. More recent event-stream approaches have further demonstrated the feasibility of directly modeling irregular clinical observations ^14^. Fixed-window representations nevertheless remain widely used because they provide a regular feature space for conventional sequence models; for example, established ICU benchmarks construct prediction inputs on an hourly grid ^23^. Here, we compared event-stream and fixed-window representations for postoperative prediction and examined how the retained ordering of individual events within 6-hour windows shaped STREAM’s risk estimates. In our experiments, STREAM and the fixed-window Time-series Transformer used closely aligned prediction objectives and attention structures. STREAM achieved higher postoperative dynamic discrimination, and within-window permutation experiments showed that reordering the same events within the same 6-hour interval could substantially alter the resulting prediction. The performance comparison suggests potential value in preserving event-level structure, while the permutation analysis shows that STREAM uses within-window ordering that fixed-window aggregation cannot retain. The ablation results further suggest that multiple components of the event-stream representation contributed to performance, with the dedicated *[BASE]* state playing a particularly important role: replacing it with the surgery token produced the largest architecture-level decrease in postoperative macro-AUPRC. Taken together, these findings suggest that the advantage of STREAM arises from the broader organization of longitudinal information at the event level, including event identity, local ordering, and maintenance of a structured clinical state.

The perioperative setting provides a clinically meaningful structure for this representation. PO-AKI remains an important complication of noncardiac surgery and is associated with both short- and long-term adverse outcomes ^1^. Surgical episodes also have a clear temporal anchor separating the preoperative state from postoperative course. In our cohort, EHR event density and first coded PO-AKI diagnoses were concentrated in the early postoperative period, followed by progressively sparser observations and fewer episodes that were still free of coded PO-AKI but subsequently received a diagnosis by postoperative day 30. STREAM was designed to reflect this trajectory: the dedicated *[BASE]* representation summarizes information available at the index surgery, and subsequent postoperative events update the risk state as new information arrives. This alignment between model state and successive perioperative information states may be particularly useful in longitudinal settings organized around a clinically meaningful anchor event followed by irregular, evolving observations.

Calibration provided a complementary view of model performance by assessing agreement between predicted probabilities and observed outcome frequencies ^24,25^. STREAM showed closer agreement between predicted probabilities and observed coded PO-AKI frequencies across postoperative time points, whereas the fixed-window time-series models showed greater overprediction during the early postoperative period. These findings indicate that STREAM’s performance advantage extended beyond discrimination to probabilistic risk estimation across time. However, calibration is population- and setting-dependent, and miscalibration was evident, particularly in sparsely populated high-risk regions. External validation and site-specific recalibration would therefore be necessary before clinical implementation.

Interpretability is another key consideration for longitudinal clinical prediction because model estimates depend on both the information recorded and when it becomes available.

Conventional post-hoc explanation methods such as LIME, SHAP, and Integrated Gradients primarily attribute model outputs to input features, while approaches including RETAIN, TimeSHAP, and GRU-DA provide interpretations at the visit, time-step, or feature level ^26–31^ . Event-stream modeling also allows changes in estimated risk to be examined as individual events enter the record. As shown in our STREAM analyses, newly recorded events prompted updates that could increase or decrease model-estimated risk. Within-window permutation analysis further showed that predictions were more sensitive to event ordering in event-dense windows. At a broader temporal scale, grouped deletion identified reliance on repeated clinical codes and showed strong agreement with event-arrival attribution among overlapping codes (Spearman ρ = 0.822; 91.9% directional agreement). Age and surgery substitution further characterized how surgery-index estimates responded to structured baseline factors. For events sharing a timestamp, averaging attributions across sampled orders accounted for their unresolved ordering. Together, these findings illustrate how an event-stream representation can support interpretation of evolving model estimates as clinical information accumulates.

A further potential advantage of event-stream modeling is representational portability. Structured clinical trajectories share an important representational property with natural language: both can be expressed as sequences of discrete elements whose meaning depends on surrounding context and order. This principle has motivated prior Transformer-based EHR models that adapt contextual sequence modeling to clinical records ^18,32^. STREAM extends this formulation by representing diagnoses, medications, laboratory measurements, and procedures within a common event sequence while encoding temporal and numerical information alongside event identity. Such a representation provides a route toward reducing dependence on manually constructed, site-specific feature matrices. Standardized data models and vocabularies, including those developed within OHDSI, could further support this direction by mapping heterogeneous local records to shared clinical concepts ^33^. Future work should therefore evaluate whether event-stream models trained on standardized clinical tokens can transfer across institutions, clinical domains, and prediction tasks, while accounting for differences in case mix, coding practices, workflows, measurement frequency, and medication formularies.

Several limitations should be considered when interpreting these findings. First, this retrospective study was conducted within a single health system, and external validation is needed to assess transferability. Second, PO-AKI was defined using diagnosis codes and therefore reflects clinically documented coded AKI, with potential under capture of physiologic KDIGO-defined kidney injury; death was not modeled as a competing event. Third, EHR event streams reflect both patient state and patterns of clinical observation, and the recorded temporal resolution does not always resolve the ordering of events sharing the same timestamp.

Although permutation averaging reduced dependence on this ambiguity in the attribution analyses, unresolved temporal granularity remains inherent to routinely collected EHR data. Fourth, the number of positive episodes remaining in the postoperative risk set decreases at later time points, making late-period discrimination and calibration estimates less stable. Finally, this study evaluated dynamic prediction in an offline retrospective setting and did not assess a deployed clinical alerting workflow. Appropriate alert thresholds, false-alert burden, workflow integration, site-specific recalibration, and prospective effects on clinical decision-making remain to be established before clinical implementation.

To conclude, we developed STREAM, a perioperative event-stream Transformer that supports surgery-index risk estimation and event-by-event postoperative updating from irregular EHR data. In a large surgical cohort, STREAM improved surgery-index and postoperative dynamic prediction of coded PO-AKI and showed more consistent calibration across postoperative time points. Event-arrival attribution and within-window permutation analyses further showed that model predictions changed with newly arriving clinical information and were sensitive to fine-grained event ordering. These findings support event-stream modeling as a promising approach for preserving event-level temporal structure while enabling dynamically updated and interpretable prediction from longitudinal EHR data.

## Methods

### Ethical Approval

This study was approved by the University of Texas Institutional Review Board (IRB) (IRB Number: HSC-SBMI-24-0403). The requirement for written informed consent was waived by the ethics committee due to the retrospective, de-identified nature of the study.

### Data Source and OMOP Transformation

The study used EHR data from the Memorial Hermann Health System in Houston, Texas, collected between January 2014 and April 2024. Raw clinical data from the Cerner EHR system were transformed into the Observational Medical Outcomes Partnership Common Data Model (OMOP CDM, version 5.4) through an institutional extract-transform-load (ETL) pipeline. Patient records were de-identified and deduplicated using an institutional Master Patient Index to support longitudinal linkage across encounters.

### Study Cohort Construction

From 768,613 OMOP CDM procedure records involving 689,015 patients, we selected the 200 most frequent procedure types (602,871 records; 516,738 patients) and mapped them to SNOMED CT concepts. We sequentially excluded noninvasive procedures (n = 379,195), cardiac surgeries (n = 4,995), procedures in patients younger than 18 years (n = 10,934), procedures with an AKI diagnosis recorded in the preceding year (n = 6,435), and procedures in patients with unknown sex (OMOP concept ID 0; n = 34,234), as summarized in **Figure 1A**. Eligible procedures were categorized as abdominal, intracranial, thoracic, spinal, joint, or skin surgery. Each eligible surgery defined a surgical episode, with a perioperative observation window extending from 7 days before surgery through postoperative day 30. Episode eligibility was determined independently of subsequent coded PO-AKI status.

### Outcome Definition

The primary outcome was coded PO-AKI within 30 days after the index surgery, identified using ICD-9-CM codes beginning with 584 or ICD-10-CM codes beginning with N17. The first qualifying postoperative diagnosis defined the PO-AKI event time. For episodes with coded PO-AKI, prediction inputs were restricted to information available before this event; otherwise, follow-up extended through postoperative day 30.

### Event Stream Construction

Each eligible noncardiac surgical procedure was treated as an index surgery and anchored at *t*_0_ = 0. For each surgical episode, the perioperative observation window extended from 7 days before the index surgery through 30 days after surgery. For episodes with coded PO-AKI, the first qualified postoperative AKI diagnosis defined the endpoint, and the event stream was truncated at that point to prevent post-outcome information from entering the model. Within each observation window, EHR data were extracted from five clinical domains: demographics, conditions, medications, laboratory tests, and the index surgery. Each record was retained as an individual clinical event and positioned according to its time relative to the index surgery, yielding a chronologically ordered sequence of heterogeneous, irregularly spaced events.

### Event Vocabulary and Tokenization Strategy

A unified event vocabulary was constructed to map heterogeneous EHR records into discrete clinical event tokens (**Supplementary Table S7**). The complete vocabulary, including retained clinical codes and their corresponding token definitions, is provided in **Supplementary Data 1**.

#### Demographics

Age, sex, and race were represented as baseline demographic events and positioned at the beginning of the observation window (day ™7). Age was represented by a categorical event identity together with its numerical value, whereas sex and race were represented as categorical event codes.

#### Conditions

Diagnosis records were mapped from ICD-9-CM and ICD-10-CM codes to Clinical Classifications Software (CCS) categories defined by the Healthcare Cost and Utilization Project (HCUP) ^34^. This mapping reduced coding heterogeneity while preserving clinically meaningful diagnostic groups.

#### Medications

Medication records were normalized to ingredient-level RxNorm concept unique identifiers (RxCUIs). Ingredients accounting for the top 90% of medication event frequency were retained as individual vocabulary entries, and less frequent ingredients were mapped to a generic *MED_OTHERS* token. Medication start and end information was retained as an event-level boundary attribute and subsequently encoded by the medication-boundary embedding.

#### Laboratory Tests

21 commonly measured and renal-function-related laboratory tests were retained as individual event types, whereas all remaining laboratory measurements were mapped to *LAB_OTHERS*. After unit harmonization within each laboratory test, the observed numerical measurement was retained as a scalar event value.

#### Index Surgery

The performed surgery was represented using its mapped SNOMED CT procedure concept.

### The Event Stream Representation and Model Architecture

Figure 6. provides an overview of the event-stream representation and STREAM architecture. Following tokenization, each surgical episode was aligned to the recorded completion timestamp of the index surgery *t*_0_ = 0. The available OMOP CDM records did not reliably identify whether individual events occurred intraoperatively, so no separate intraoperative segment was defined. Other events were positioned by their recorded event timestamps relative to this anchor. Events before the anchor, together with the index surgery token, formed the pre-index segment; events after it formed the postoperative segment. The pre-index segment could therefore include intraoperative records. STREAM processed the resulting chronological event stream as follows:

**Figure 6.**
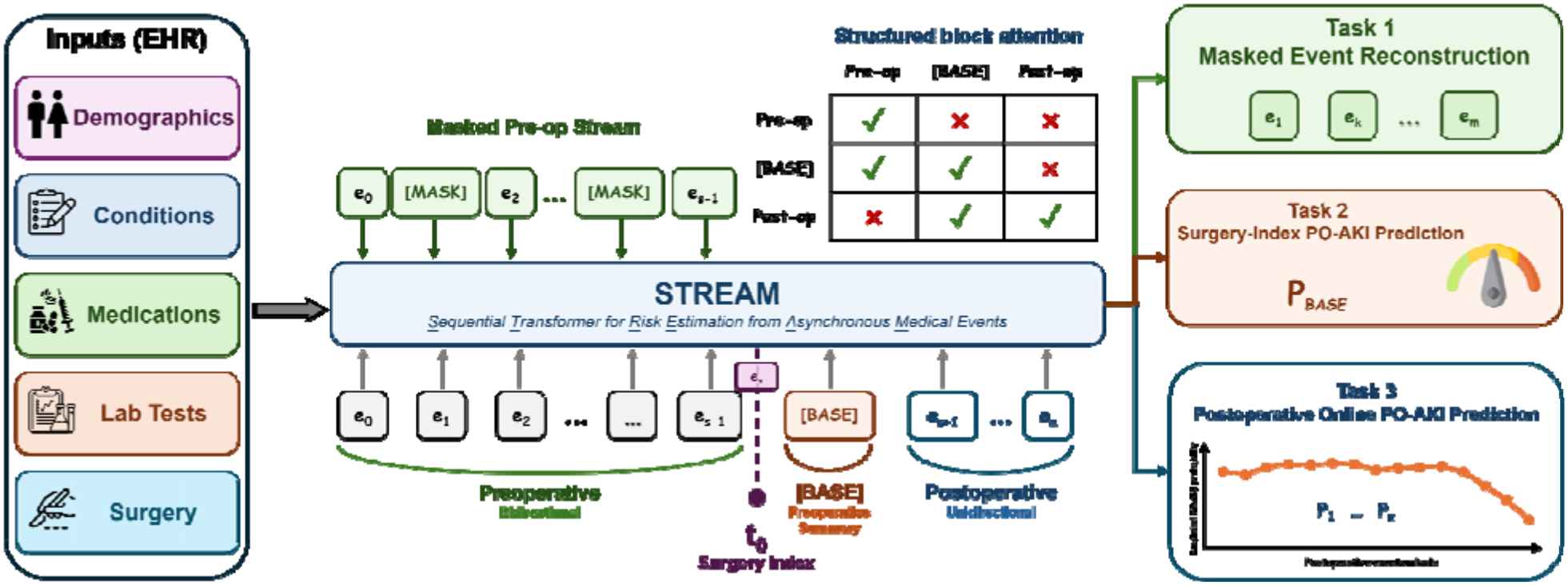
Architecture of STREAM (Sequential Transformer for Risk Estimation from Asynchronous Medical Events). Heterogeneous perioperative EHR events are represented as an irregular chronological sequence anchored at the index surgery. STREAM uses structured block attention with bidirectional encoding of preoperative events, a dedicated representation summarizing the patient state available at surgery, and unidirectional processing of postoperative events for online risk updating. The shared encoder is jointly trained for preoperative masked event code reconstruction, surgery-index PO-AKI prediction, and postoperative online PO-AKI prediction.

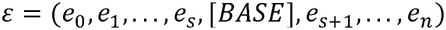

where *e*_O_,…, *e*_*s −1*_each denotes a pre-index event, *e*_*s*_ denotes the index surgery at *t*_0_ = 0, and *e*_*s+1*_,…, *e*_*n*_ each denotes a postoperative event. A learnable special token *[BASE]* was inserted immediately after the surgery event to summarize the patient state available at the surgery-index.

Each clinical event *e*_*i*_ was defined as:

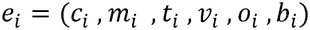

Where *c*_*i*_ denotes the clinical event code, *m*_*i*_ the event modality, *t*_*i*_ the relative time from the index surgery, *v*_*i*_ an optional scalar value, *o*_*i*_ an indicator of whether a numerical value was observed, and *b*_*i*_ the medication boundary type. For medication events, *b*_*i*_ indicated medication start or end; non-medication events were assigned the null boundary type.

Each event was mapped to a dense representation by summing learnable embeddings and continuous-feature projections:

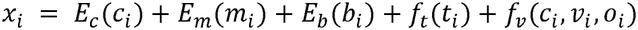

Here, *E*_*C*_ and *E*_*m*_ are learnable embeddings for clinical codes and event modalities, respectively. Medication boundary information was encoded by *E*_*b*_ with three states: none, medication start, and medication end; the null state was mapped to a zero vector. *f*_*t*_ is a learnable projection of relative time from the index surgery. Numerical values were present only for age and laboratory events. Age and laboratory values were normalized separately using statistics estimated from the corresponding training fold and projected through dedicated MLPs within *f*_*v*_ ; events without an observed numerical value received no value contribution. All embedding and projection components were optimized jointly with the Transformer encoder. Variable-length event streams were padded within each batch, with padding positions excluded from attention and loss computation.

To support surgery-index prediction and postoperative updating, STREAM used a structured block attention mask. Pre-index events, including the index surgery, attended bidirectionally within the pre-index segment. The *[BASE]* token attended to all preoperative events and itself, allowing it to summarize information available at surgery. Postoperative events were processed using unidirectional, past-only attention: each event attended to *[BASE]* and postoperative events observed up to and including the current event, while future postoperative events were masked. Thus, each postoperative prediction used only information available at that point in clinical time.

The Transformer generated hidden representations

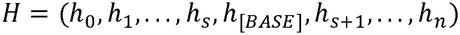

The hidden state *h*_[*BASE*]_ was used to estimate surgery-index PO-AKI risk, whereas each postoperative hidden state *h*_*i*_ generated an updated online risk estimate as new clinical events entered the stream.

### Multi-task Training Objectives

STREAM was optimized end-to-end using three complementary objectives:

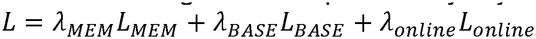

The postoperative online prediction objective served as the primary clinical objective, with surgery-index prediction and masked event reconstruction providing additional supervision.

***Task 1: Preoperative Masked Event Code Reconstruction (****L*_***MEM***_)

During training, 15% of eligible preoperative event codes were randomly replaced with a [*MASK*] token. The model was trained to reconstruct the original clinical code from the surrounding preoperative context. For masked laboratory events, the associated numerical value and observation indicator were also masked for reconstruction. For the set of masked pre-index positions *M*_*pre*_ , the reconstruction loss was defined as

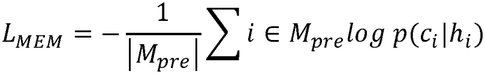

This objective encouraged the encoder to learn contextual relationships among heterogeneous preoperative clinical events.

***Task 2: Surgery-index PO-AKI Prediction (****L*_***BASE***_)

The *[BASE]* hidden representation was used to estimate PO-AKI risk from information available at the surgery-index:

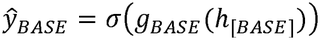

where *g*_*BASE*_ denotes the baseline prediction head. The corresponding loss was

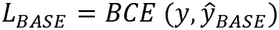

This objective explicitly trained the *[BASE]* representation for surgery-index risk estimation.

***Task 3: Postoperative Online PO-AKI Risk Prediction (****L*_***online***_)

For each postoperative event *e*_*i*_, STREAM generated an updated PO-AKI probability:

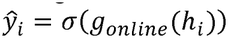

The online prediction loss was calculated over valid postoperative event positions:

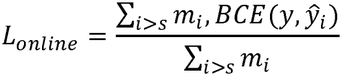

where *m*_*i*_ indicates whether position corresponds to a valid postoperative event. Because postoperative attention was unidirectional and future events were masked, each ŷ_i_ was based only on the *[BASE]* state and postoperative information available up to that event. This enabled STREAM to transition from surgery-index prediction to event-by-event postoperative risk updating within a shared architecture.

### Training Details

The cohort was split chronologically by surgery date. The earliest 80% were assigned to the development period (1 Jan. 2014–29 Aug. 2022), and later surgeries were initially reserved for temporal testing (30 Aug. 2022–13 Apr. 2024). No patient contributed surgeries to both sets. Within the development set, five-fold cross-validation was used for model development and checkpoint selection, with all episodes from the same patient assigned to the same fold to prevent patient-level information leakage. For the final held-out temporal test set, only the first eligible surgical episode from each patient was retained to ensure patient-level independence during evaluation.

All data-dependent preprocessing was estimated separately within each training fold. The event vocabulary and numerical normalization parameters, including age and laboratory-value statistics, were derived exclusively from the corresponding training data and subsequently applied to the associated validation data and held-out temporal test set. The held-out temporal test set was not used for model development, hyperparameter tuning, or checkpoint selection. For each fold, checkpoints were selected using a composite validation score prioritizing postoperative dynamic prediction:

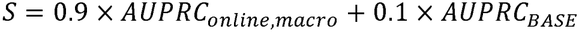

where *AUPRC*_*BASE*_ denotes surgery-index AUPRC and AUPRC_*online, macro*_ denotes the unweighted mean AUPRC across valid postoperative 6-hour intervals.

The best checkpoint from each fold was independently evaluated on the same held-out temporal test set, and performance was summarized as the mean (minimum, maximum) across the five fold-specific models. Hyperparameters were tuned within the development data, and the final STREAM configuration is provided in **Supplementary Table S8**.

### Comparative Models

STREAM was compared with static and fixed-window time-series models using the same development and held-out temporal test sets. All models were trained to predict the same 30-day coded PO-AKI outcome, with static models evaluated at the surgery-index and sequence models evaluated at both the surgery-index and during postoperative follow-up.

A common 548-dimensional feature space was constructed from demographics, surgical categories, CCS diagnosis groups, laboratory indicators and values, and medication features. For fixed-window models, the perioperative observation window was discretized into a regular 6-hour grid. The 7-day preoperative period yielded 28 intervals, and the 30-day postoperative period yielded 120 intervals. A dedicated surgery-index baseline position was inserted between the preoperative and postoperative segments, resulting in a sequence of 149 positions. Each position was represented by the corresponding 548-dimensional feature vector.

### Static Models

Static models were used to benchmark surgery-index PO-AKI prediction using information recorded by completion of the index surgery. Categorical features were represented as binary indicators, and the most recent available preoperative laboratory measurement was used for each laboratory feature.

#### Logistic regression

An L2-regularized logistic regression model served as the linear baseline. **XGBoost**. A gradient-boosted tree classifier was used as a nonlinear static baseline, with hyperparameters selected by grid search (**Supplementary Table S9**).

### Time-series Models

Three fixed-window time-series models were used to benchmark surgery-index and postoperative dynamic PO-AKI prediction. Pseudocode for all three models is provided in **Supplementary Algorithm 1**.

#### LSTM

A unidirectional long short-term memory (LSTM) network processed the 149-position sequence after projection to a 128-dimensional hidden representation. Separate prediction heads generated surgery-index risk from the baseline position and updated postoperative risk at subsequent 6-hour intervals.

#### GRU-D

GRU-D ^22^ was used to explicitly model missingness and irregular observation patterns through trainable decay mechanisms. Missing features were dynamically decayed toward the training-fold population mean according to the elapsed time since their most recent observation, while the observation mask and time-since-last-observation information were incorporated into the recurrent updates.

#### Time-series Transformer

A two-layer Transformer encoder with learned positional embeddings was applied to the same 149-position sequence. Its attention structure paralleled that of STREAM: pre-index positions attended bidirectionally within the pre-index segment; the surgery-index baseline position summarized information recorded by surgery completion; and each postoperative position attended only to the baseline representation and postoperative positions up to the current interval.

For the LSTM and Time-series Transformer, missing numerical values were handled using last observation carried forward (LOCF), followed by population-mean imputation when no prior observation was available. All imputation statistics were estimated from the corresponding training fold. The three sequence baselines were trained using the same surgery-index and postoperative prediction objectives and selected using the same composite validation criterion as STREAM.

### Predictive Performance Evaluation

Performance was evaluated at both the surgery-index and postoperative dynamic stages using AUROC, AUPRC, accuracy, precision, sensitivity (recall), specificity, and F1-score. Binary classification metrics were calculated using a probability threshold of 0.5. AUROC and AUPRC were reported as the primary discrimination metrics in the Results section, with complete metrics provided in the Supplementary Tables.

For surgery-index evaluation, STREAM was compared with both static and fixed-window time-series models on the held-out temporal test set using the same 30-day coded PO-AKI prediction target.

For postoperative dynamic evaluation, the sequence models were evaluated on a common 6-hour grid comprising 120 intervals over the 30-day postoperative period. Fixed-window models generated one prediction per interval, whereas STREAM generated predictions whenever new clinical events occurred. STREAM predictions were aligned to the common grid using the most recent prediction available by each evaluation time point; the *[BASE]* prediction was carried forward until the first postoperative event when necessary.

Performance was calculated separately within each valid postoperative interval. For each metric, the postoperative macro-average was defined as the unweighted mean of the corresponding interval-specific values across all valid 6-hour intervals; intervals in which the metric was undefined were excluded. AUPRC was prespecified as the primary metric for discrimination across successive 6-hour postoperative intervals because coded PO-AKI was an imbalanced outcome.

### Timepoint-Specific Calibration Evaluation of PO-AKI Risk Prediction

Calibration assesses whether predicted probabilities correspond to observed event frequencies. In practice, episodes are grouped according to predicted PO-AKI probability, and the mean predicted probability is compared with the observed event rate within each group. Predictions were divided into 10 equal-frequency groups, and calibration curves plotted mean predicted risk against the corresponding observed PO-AKI proportion. A well-calibrated model should produce predicted risks that closely match observed outcome frequencies; therefore, closer agreement with the 45° identity line indicates better calibration ^24,25^.

Calibration was evaluated at 0, 6, 12, 24, 48, and 168 hours after the index surgery. At each time point, the analysis included episodes that remained free of coded PO-AKI, with the outcome defined as subsequent coded PO-AKI through postoperative day 30. For STREAM, the most recent prediction available at or before each evaluation time point was used: the *[BASE]* prediction at 0 hours and the latest postoperative event-level prediction thereafter. If no postoperative event had occurred, the *[BASE]* prediction was carried forward. Fixed-window time-series models were evaluated using the most recent available 6-hour prediction, whereas static models were evaluated only at the surgery-index.

Calibration was evaluated separately for each of the five fold-specific models, and the mean calibration curve across folds was displayed. Predicted-risk distributions were shown beneath each calibration panel. Brier score, Brier skill score, calibration intercept, and calibration slope were additionally reported as quantitative calibration measures in the Supplementary material. The Brier skill score was calculated relative to a reference model assigning the observed outcome prevalence at each evaluation time point to all episodes. Calibration intercept and slope were estimated by regressing the binary outcome on the logit-transformed predicted probabilities, with ideal values of 0 and 1, respectively.

### Ablation Study of Model Architecture, Training Objectives, and Inputs

To quantify the contributions of architectural components, training objectives, and input modalities, we trained a series of single-factor and joint ablation variants of STREAM. All variants followed the same training and checkpoint-selection protocol described above, except where the corresponding component or objective was explicitly ablated. For the no-baseline-loss variant (A4), checkpoint selection was based solely on postoperative macro-AUPRC. Architecture and training-objective ablations included removal of continuous time encoding (A1); replacement of the structured block attention mask with unidirectional, past-only attention throughout the sequence (A2); removal of the dedicated *[BASE]* token, with the index-surgery token used as the surgery-index representation (A3); removal of the surgery-index prediction loss (λ_BASE_ = 0; A4); and removal of the MEM auxiliary objective (λ_MEM_ = 0; A5). Input ablations removed medication events (I1), laboratory-test events (I2), condition events (I3), or surgery identity by replacing the surgery-specific token with a generic surgery token (I4). We additionally evaluated a joint A3+I4 variant in which the dedicated *[BASE]* token was removed and the generic surgery token served as the surgery-index representation, allowing the contribution of a dedicated baseline representation to be examined independently of surgery identity. Each variant was trained independently and evaluated on the common held-out temporal test set. The primary ablation outcomes reported in the main manuscript were surgery-index AUPRC and postoperative macro 6-hour AUPRC, with complete performance metrics provided in the Supplementary material.

### Interpretability and Temporal Information Use

#### Event-Arrival Attribution

The event-stream architecture provides a natural event-level view of how STREAM updates PO-AKI risk as new postoperative information becomes available. We therefore performed event-arrival attribution to quantify the change in the online prediction associated with each newly observed clinical event.

For postoperative event *e*_*i*_, let *z*_*i*_ denote the PO-AKI logit produced by the online prediction head after incorporating that event. Event-arrival attribution was defined as 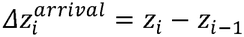 , where z_*i−1*_, denotes the immediately preceding online state. For the first postoperative event, the *[BASE]* prediction served as the preceding state. A positive 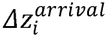 indicates an increase in model-estimated PO-AKI risk after the event is incorporated, whereas a negative value indicates a decrease.

### Order-Robust Attribution for Simultaneous Events

For events sharing an identical recorded timestamp, the EHR does not provide sufficient temporal resolution to determine their relative order. For each simultaneous-event group *G*_*t*_ = {*e*_*1*_,*…, e*_*k*_}, we therefore sampled up to *R*= 20 within-group permutations and recalculated the marginal logit contribution of each event under each ordering. The attribution assigned to an event was the mean of its marginal logit contributions across the sampled permutations, providing an order-robust estimate under timestamp-level ordering uncertainty.

Event-arrival attribution was evaluated on the full held-out temporal test set (*n* = 23,538) independently for each of the five fold-specific STREAM models. Clinical codes were retained if they were observed in at least 50 patients in at least one fold. A code-level attribution was considered replicated when its mean contribution had the same sign across all five models.

Primary code-level analyses were restricted to the first 7 postoperative days, when event density and the number of episodes remaining at risk were greatest.

We in addition provide an illustrative patient-level case study of event-arrival attribution. One coded PO-AKI-positive episode and one episode without coded PO-AKI were selected, and sequential postoperative predictions were aligned with the clinical events entering the stream. For simultaneous events, the permutation-averaged attribution described above was used. For visualization, model logits were transformed to predicted probabilities, *p*_i_ =*σ*(*z*_*i*_), and the resulting probability trajectory and event-associated updates were displayed over postoperative time. As a robustness analysis, events within identical timestamps were randomly shuffled five times while all timestamps and other sequence information were preserved, and the resulting absolute changes in model logit were quantified.

### Within Window Permutation Analysis

To assess whether STREAM used fine-grained event ordering within the fixed 6-hour evaluation windows, we performed a within-window permutation analysis. For each eligible postoperative window, the clinical event content and original timestamp were preserved, while events were randomly reassigned across the available temporal positions. Thus, each permuted sequence contained the same events and timestamp distribution as the original window but differed in the assignment of events to earlier or later positions. We sampled up to *R*= 20 random permutations per window and compared the end-of-window PO-AKI logit with that from the original event sequence. Order sensitivity was defined as the mean absolute change in logit:

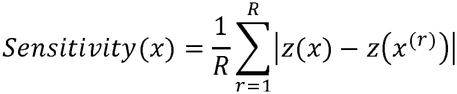

where *z(x)*denotes the end-of-window logit for the original sequence and *x*^(*r*)^ denotes the *r*-th within-window permutation. Sensitivity was summarized by postoperative time period and within-window event count *k* to examine whether order dependence varied with postoperative timing and event density.

As an illustrative test, we additionally reversed the event order within a representative first 6-hour postoperative window while retaining the same event content and temporal positions, and compared the resulting risk trajectory and end-of-window prediction with those of the original sequence.

### Supplementary Perturbation-Based Attribution Analyses

To complement the event-arrival and order-sensitivity analyses, we performed three perturbation-based analyses to characterize STREAM’s reliance on clinical inputs: temporal grouped deletion, index-surgery substitution, and age-value substitution.

### Temporal Grouped Deletion

To characterize how clinical events contributed to model-estimated PO-AKI risk, we used a perturbation-based attribution framework. Perturbation methods estimate input importance by modifying or removing selected inputs and measuring the resulting change in model output ^35^. For irregular EHR event streams, single-token perturbation may inadequately represent recurring clinical signals, and replacement with a placeholder does not reproduce true event absence when the placeholder remains visible to the attention mechanism ^36^. We therefore used a temporal grouped-deletion strategy tailored to the event-stream representation (**Supplementary Figure S4**).

Each attribution unit was defined as: *g*= *(m, c, b)* , where *m* denotes the event modality, *c* the clinical code, and *b* a predefined time window relative to the index surgery. Preoperative events were grouped into [7*d*, 48*h*) and [48*h*, 0), and postoperative events into [0, 6*h*), [6, 24*h*) ,[24*h*, 72*h*) ,[72*h*,7*d*). For a given group (*g*), all occurrences of the corresponding clinical code within the specified time window were physically removed from the input sequence, after which the sequence and attention mask were rebuilt. The attribution of event group *g* was defined as the change in predicted PO-AKI log-odds between the original and perturbed event streams. Let *z(x)* denote the PO-AKI logit for the original event stream *x* , and *x*_−g_ the stream after deleting group *g* . The attribution score was:

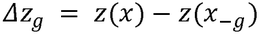

A positive , Δ*z*_*g*_ indicates that the event group increased the model-estimated PO-AKI logit relative to its deletion, whereas a negative value indicates a downward contribution. Attribution was quantified on the logit scale to reduce compression of effect magnitude near probability boundaries.

Surgery-index attribution was evaluated at the *[BASE]* representation, whereas postoperative attribution was evaluated at prediction positions occurring at or after the corresponding event group. Postoperative attribution was restricted to the first 168 hours (7 days) after surgery to reduce instability from increasingly sparse late postoperative risk sets.

Within each surgical episode, attribution values for a given group (*m, c, b*) were averaged across eligible query positions to obtain one episode-level contribution estimate. Groups represented by fewer than 25 distinct patients were excluded. Attribution analysis was performed on the full held-out temporal test set (n=23,538) and repeated independently across the five fold-specific models. For each group, we calculated the mean attribution across folds, the number of folds with concordant attribution direction, and a patient-level bootstrap 95% confidence interval (CI) based on 2,000 resamples. A contribution was considered replicated when all five models agreed in sign, and the bootstrap 95% CI excluded zero.

### Index Surgery Substitution

The index surgery required a separate attribution strategy because each surgical episode contained exactly one surgery token at *t*_0_ = 0. Deleting this token would create an input without a defining surgical procedure and yield a difficult-to-interpret perturbation. We therefore used surgery substitution.

For each episode in the held-out temporal test set, all non-surgery events and covariates were held fixed while the index surgery token was sequentially replaced with each eligible SNOMED CT procedure code. STREAM was then rescored to estimate the mean surgery-index logit associated with each substituted surgery:

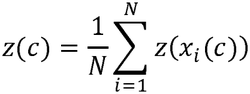

Where *x*_i_ (*c*) denotes the event stream for episode *i* with the index surgery replaced by surgery *c*, and *z*(·) denotes the corresponding STREAM surgery-index logit. This analysis quantified how surgery identity altered the model prediction while the remaining patient context was held fixed.

surgery-specific effects were expressed relative to a reference surgery:

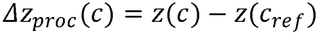

The reference surgery was selected as the surgery with the median z (*c*) across all eligible surgeries; in this analysis, it was minimally invasive decompression of lumbar spine (SNOMED CT 830198007). Surgery substitution was performed independently for each of the five fold-specific STREAM models, and surgery-specific logits were averaged across folds before relative effects were calculated. The analysis included the full held-out temporal test set (*n* = 23,538) and 75 eligible surgery types.

As an empirical consistency analysis, surgery-level substitution effects were compared with the observed coded PO-AKI incidence among episodes in which each surgery was actually performed. Spearman rank correlation was calculated between Δ*z*_*proc*_ (*c*) and the observed surgery-specific coded PO-AKI incidence to assess concordance between model-derived surgery effects and empirical surgery-level risk patterns.

### Age Value Substitution

Age was evaluated separately because the AGE token contains a continuous numerical value. Deleting the token would quantify the effect of removing age information but would not characterize the direction or shape of the age–risk relationship. We therefore performed a within-episode value-substitution analysis in which age was varied from 20 to 90 years in 5-year increments while all other event-stream inputs were held fixed. Using 48 years, corresponding to the cohort median age, as the reference, the age-substitution effect was defined as *Δ z*_*age*_ *(a) = z(x*_*a*_*) − z(x*_*48*_) , where *x*_*a*_ denotes the event stream with age set to *a*. Age-response curves were averaged across the evaluated episodes and the five fold-specific STREAM models to characterize how age information influenced model predictions.

These perturbation-based estimates were interpreted as measures of model reliance under input modification, not as causal effects.

## Data Availability

All data produced in the present study are available upon reasonable request to the authors.

https://github.com/Frank-LSY/STREAM-PO-AKI

## Statistics and Reproducibility

All eligible surgical episodes within the study period were included; no formal sample-size calculation was performed. Model development used five-fold cross-validation with patient-level fold assignment, and each fold-specific model was evaluated on the same held-out temporal test set. Predictive performance was summarized as the mean (minimum, maximum) across folds. Postoperative dynamic metrics were calculated within valid 6-hour intervals and macro-averaged across intervals, with AUPRC prespecified as the primary metric. Threshold-dependent metrics used a probability threshold of 0.5. Episode-level bootstrap resampling with 2,000 resamples was used for grouped-deletion attribution confidence intervals, and 95% confidence intervals for within-window order sensitivity were estimated using 2,000 window-level bootstrap resamples. Spearman rank correlation was used for attribution concordance analyses. Randomization and investigator blinding were not applicable to this retrospective computational study.

## Data Availability

The datasets generated and/or analyzed during the current study are not publicly available due to patient privacy and confidentiality restrictions but are available from the corresponding author on reasonable request.

## Code Availability

The underlying code for model, analysis, result tables and figures for this study is available in STREAM-PO-AKI and can be accessed via this link https://github.com/Frank-LSY/STREAM-PO-AKI.

## Acknowledgements

This study was funded by National Human Genome Research Institute (3R01HG012748-02S2), Cancer Prevention and Research Institute of Texas Established Investigator Award (RR230020), and the National Institute of Aging (R01AG072799-03). The funders played no role in study design, data collection, analysis and interpretation of data, or the writing of this manuscript.

## Author Contributions

SL: study design, cohort construction, model development and training, data analysis, interpretation of results, and drafting of the manuscript.

XR: project coordination, cohort definition, methodological advice, and critical revision of the manuscript.

JA, AW: preliminary data curation and cohort construction.

LW, XW: data interpretation and critical revision of the manuscript.

YL: clinical input on cohort definition, interpretation of findings, and critical revision of the manuscript.

HL: project supervision, and critical revision of the manuscript. JJ, XL, YH: literature review and revision of the manuscript.

All authors reviewed and approved the final manuscript.

