## Supplementary Tables and Figures for "STREAM Sequential Transformer for Risk Estimation from Asynchronous Medical Events for Dynamic Postoperative AKI Prediction"

**Overview**

This Supplementary Information provides model specification tables, complete predictive performance metrics, quantitative calibration results, detailed ablation results, and complementary interpretation analyses for STREAM. Population-level event-arrival attribution and within-window event-order sensitivity are presented in the main manuscript, while the supplementary material provides additional quantitative results and representative patient-level analyses.

**Complementary predictive performance**

The main manuscript reports AUROC and AUPRC as the primary discrimination metrics. **Supplementary Tables S1 and S2** provide the complete surgery-index and postoperative dynamic performance metrics, respectively, including accuracy, specificity, sensitivity, precision, F1-score, AUROC, and AUPRC. Values are reported as percentages and summarized as mean (minimum, maximum) across the five fold-specific models evaluated on the same held-out temporal test set.

| **Supplementary Table S1. Complete surgery-index predictive performance.** | | | | | | | | |
| --- | --- | --- | --- | --- | --- | --- | --- | --- |
| **Model Type** | **Model** | **AUROC** | **AUPRC** | **Accuracy** | **Specificity** | **Sensitivity** | **Precision** | **F1** |
| Static | Logistic Regression | 91.05 (91.02, 91.07) | 61.17 (61.08, 61.26) | 82.01 (81.72, 82.21) | 81.82 (81.47, 82.08) | 83.71 (83.34, 83.93) | 34.04 (33.66, 34.25) | 48.39 (48.05, 48.60) |
|  | XGBoost | 90.69 (90.57, 90.88) | 60.47 (60.24, 60.68) | 85.53 (85.18, 85.81) | 86.38 (85.92, 86.80) | 77.89 (76.97, 78.57) | 39.05 (38.47, 39.50) | 52.02 (51.65, 52.24) |
| Fixed-window Time Series | LSTM | 92.42 (92.21, 92.56) | 67.76 (66.61, 68.34) | 93.47 (93.32, 93.62) | 97.93 (97.66, 98.13) | 53.59 (50.44, 55.93) | 74.46 (72.78, 75.86) | 62.29 (60.36, 63.25) |
|  | GRU-D | 92.63 (92.51, 92.83) | 69.48 (69.04, 70.28) | 93.58 (93.47, 93.70) | 97.93 (97.81, 98.07) | 54.78 (53.86, 55.50) | 74.78 (73.67, 76.04) | 63.23 (62.72, 63.82) |
|  | Time-series Transformer | 93.33 (93.27, 93.41) | 72.88 (72.72, 73.07) | 94.35 (94.32, 94.39) | 98.19 (98.05, 98.27) | 60.08 (59.05, 61.07) | 78.80 (77.81, 79.32) | 68.18 (67.70, 68.43) |
| Event Stream | STREAM | 94.19 (94.02, 94.32) | 77.14 (76.70, 77.73) | 94.95 (94.74, 95.14) | 98.36 (97.96, 98.58) | 64.52 (62.67, 66.04) | 81.59 (78.38, 83.42) | 72.03 (71.25, 72.77) |

| **Supplementary Table S2. Complete postoperative dynamic performance using macro-averaged 6-hour metrics.** | | | | | | | | |
| --- | --- | --- | --- | --- | --- | --- | --- | --- |
| **Model Type** | **Model** | **AUROC** | **AUPRC** | **Accuracy** | **Specificity** | **Sensitivity** | **Precision** | **F1** |
| Fixed-window Time Series | LSTM | 93.24 (92.79, 93.59) | 44.49 (41.97, 46.35) | 97.09 (96.90, 97.24) | 98.12 (97.90, 98.32) | 47.76 (45.58, 49.58) | 47.44 (43.22, 51.88) | 45.60 (44.35, 46.96) |
|  | GRU-D | 94.23 (93.98, 94.39) | 54.70 (53.37, 55.41) | 97.57 (97.41, 97.62) | 98.52 (98.31, 98.59) | 56.72 (53.87, 59.56) | 54.35 (50.77, 55.76) | 54.25 (53.08, 55.61) |
|  | Time-series Transformer | 94.76 (94.59, 94.86) | 52.45 (50.72, 53.85) | 97.70 (97.66, 97.73) | 98.62 (98.58, 98.68) | 54.97 (54.48, 55.31) | 54.33 (53.10, 55.48) | 53.24 (52.69, 54.09) |
| Event Stream | STREAM | 94.82 (94.72, 94.96) | 58.86 (58.06, 60.23) | 98.30 (98.12, 98.44) | 99.17 (99.03, 99.34) | 63.86 (60.28, 66.16) | 61.64 (59.06, 65.26) | 61.72 (59.39, 63.12) |

**Quantitative Calibration Results**

Calibration curves are presented in the main manuscript. **Supplementary Table S3** provides complementary quantitative calibration results across the prespecified postoperative time points, including Brier score, Brier skill score, calibration intercept, and calibration slope.

| **Supplementary Table S3. Quantitative calibration metrics across postoperative time points.** | | | | | | | |
| --- | --- | --- | --- | --- | --- | --- | --- |
| **Time point** | **Model** | **At-risk episodes, n** | **Subsequent coded PO-AKI, n (%)** | **Brier score** | **Brier skill score** | **Calibration intercept** | **Calibration slope** |
| Baseline (0 h) | Logistic Regression | 23,538 | 2,371 (10.1) | 0.134 (0.133, 0.136) | -0.478 (-0.498, -0.466) | -2.40 (-2.42, -2.39) | 0.76 (0.75, 0.77) |
|  | XGBoost |  |  | 0.105 (0.103, 0.107) | -0.157 (-0.179, -0.135) | -1.95 (-1.97, -1.93) | 0.80 (0.78, 0.82) |
|  | LSTM |  |  | 0.049 (0.047, 0.050) | 0.463 (0.452, 0.478) | 0.00 (-0.12, 0.19) | 0.83 (0.81, 0.86) |
|  | GRU-D |  |  | 0.050 (0.049, 0.050) | 0.450 (0.444, 0.459) | -0.08 (-0.13, -0.02) | 0.90 (0.88, 0.91) |
|  | Time-series Transformer |  |  | 0.046 (0.046, 0.047) | 0.492 (0.485, 0.496) | -0.16 (-0.21, -0.11) | 0.79 (0.77, 0.80) |
|  | STREAM |  |  | 0.042 (0.041, 0.044) | 0.532 (0.514, 0.546) | -0.16 (-0.34, -0.00) | 0.68 (0.64, 0.72) |
| 6 h | LSTM | 23,247 | 2,080 (8.9) | 0.098 (0.086, 0.106) | -0.208 (-0.304, -0.050) | -2.18 (-2.35, -1.94) | 0.88 (0.83, 0.92) |
|  | GRU-D |  |  | 0.104 (0.099, 0.109) | -0.276 (-0.343, -0.214) | -2.32 (-2.39, -2.25) | 0.89 (0.84, 0.93) |
|  | Time-series Transformer |  |  | 0.111 (0.106, 0.117) | -0.365 (-0.431, -0.297) | -2.54 (-2.65, -2.45) | 0.79 (0.75, 0.83) |
|  | STREAM |  |  | 0.036 (0.034, 0.037) | 0.564 (0.551, 0.584) | -0.70 (-0.79, -0.58) | 0.65 (0.64, 0.66) |
| 12 h | LSTM | 23,164 | 1,997 (8.6) | 0.079 (0.073, 0.085) | -0.005 (-0.078, 0.071) | -1.94 (-2.06, -1.80) | 0.85 (0.79, 0.89) |
|  | GRU-D |  |  | 0.077 (0.074, 0.083) | 0.019 (-0.055, 0.061) | -1.92 (-2.05, -1.83) | 0.92 (0.88, 0.96) |
|  | Time-series Transformer |  |  | 0.067 (0.063, 0.068) | 0.154 (0.134, 0.197) | -1.91 (-1.96, -1.82) | 0.81 (0.76, 0.87) |
|  | STREAM |  |  | 0.034 (0.033, 0.035) | 0.567 (0.554, 0.585) | -0.75 (-0.86, -0.64) | 0.66 (0.64, 0.67) |
| 24 h | LSTM | 22,983 | 1,816 (7.9) | 0.057 (0.050, 0.059) | 0.223 (0.189, 0.311) | -1.52 (-1.69, -1.27) | 0.79 (0.74, 0.84) |
|  | GRU-D |  |  | 0.059 (0.057, 0.060) | 0.195 (0.172, 0.215) | -1.64 (-1.71, -1.61) | 0.84 (0.81, 0.88) |
|  | Time-series Transformer |  |  | 0.051 (0.048, 0.053) | 0.303 (0.266, 0.335) | -1.62 (-1.70, -1.53) | 0.80 (0.75, 0.84) |
|  | STREAM |  |  | 0.031 (0.030, 0.031) | 0.578 (0.567, 0.590) | -0.71 (-0.83, -0.61) | 0.68 (0.67, 0.70) |
| 48 h | LSTM | 22,781 | 1,614 (7.1) | 0.049 (0.045, 0.052) | 0.259 (0.213, 0.310) | -1.33 (-1.52, -1.19) | 0.74 (0.71, 0.77) |
|  | GRU-D |  |  | 0.043 (0.042, 0.045) | 0.340 (0.315, 0.360) | -1.27 (-1.34, -1.21) | 0.88 (0.85, 0.91) |
|  | Time-series Transformer |  |  | 0.040 (0.039, 0.041) | 0.394 (0.382, 0.405) | -1.29 (-1.31, -1.25) | 0.77 (0.74, 0.79) |
|  | STREAM |  |  | 0.028 (0.027, 0.028) | 0.578 (0.568, 0.585) | -0.66 (-0.78, -0.58) | 0.70 (0.68, 0.72) |
| 7 d | LSTM | 22,390 | 1,223 (5.5) | 0.042 (0.037, 0.044) | 0.193 (0.140, 0.286) | -1.28 (-1.42, -1.06) | 0.67 (0.63, 0.73) |
|  | GRU-D |  |  | 0.032 (0.031, 0.033) | 0.386 (0.351, 0.400) | -1.07 (-1.19, -1.00) | 0.84 (0.82, 0.87) |
|  | Time-series Transformer |  |  | 0.030 (0.029, 0.031) | 0.423 (0.406, 0.438) | -1.06 (-1.11, -1.01) | 0.77 (0.75, 0.78) |
|  | STREAM |  |  | 0.022 (0.021, 0.022) | 0.579 (0.571, 0.586) | -0.63 (-0.72, -0.53) | 0.72 (0.70, 0.75) |
| Values are the mean (minimum, maximum) across five fold-specific models evaluated on the same held-out temporal test set. Static models were evaluated only at the surgery index. | | | | | | | |

**Detailed ablation results**

The main manuscript summarizes the primary ablation results using surgery-index AUPRC and postoperative macro-AUPRC. **Supplementary Tables S4 and S5** provide the complete surgery-index and postoperative dynamic performance metrics, respectively. Medication removal produced the largest decrease across both evaluations, while condition removal also reduced performance. Architecture and training-objective ablations generally produced smaller changes in postoperative discrimination.

| **Supplementary Table S4. Complete surgery-index performance of STREAM ablation variants.** | | | | | | | | |
| --- | --- | --- | --- | --- | --- | --- | --- | --- |
| **Category** | **Ablation** | **AUROC** | **AUPRC** | **Accuracy** | **Specificity** | **Sensitivity** | **Precision** | **F1** |
| Full model | — | 94.19 ± 0.13 | 77.14 ± 0.49 | 94.95 ± 0.15 | 98.36 ± 0.28 | 64.52 ± 1.51 | 81.59 ± 2.21 | 72.03 ± 0.59 |
| Architecture | No time encoding (A1) | 94.06 ± 0.15 | 76.37 ± 0.70 | 94.73 ± 0.25 | 98.39 ± 0.50 | 62.11 ± 2.28 | 81.47 ± 4.11 | 70.38 ± 0.67 |
|  | Unidirectional attention (A2) | 94.06 ± 0.32 | 77.33 ± 0.89 | 94.93 ± 0.21 | 98.42 ± 0.37 | 63.77 ± 1.36 | 82.06 ± 3.25 | 71.72 ± 0.48 |
|  | Procedure token as baseline (A3) | 93.99 ± 0.14 | 76.91 ± 0.43 | 95.04 ± 0.08 | 98.55 ± 0.16 | 63.74 ± 1.03 | 83.13 ± 1.41 | 72.14 ± 0.41 |
| Training objective | No baseline loss (A4)^†^ | 47.84 ± 28.20 | 15.36 ± 12.15 | 48.82 ± 31.35 | 47.40 ± 38.92 | 61.50 ± 44.35 | 9.78 ± 7.49^‡^ | 16.58 ± 12.23^‡^ |
|  | No MEM loss (A5) | 94.08 ± 0.20 | 76.83 ± 1.13 | 94.89 ± 0.37 | 98.31 ± 0.56 | 64.38 ± 1.84 | 81.30 ± 4.40 | 71.77 ± 1.17 |
| Input modality | No medication events (I1) | 89.28 ± 0.17 | 52.49 ± 0.54 | 91.28 ± 0.19 | 98.20 ± 0.52 | 29.46 ± 6.28 | 65.22 ± 2.87 | 40.12 ± 6.23 |
|  | No laboratory events (I2) | 94.34 ± 0.07 | 78.31 ± 0.28 | 95.17 ± 0.10 | 98.86 ± 0.28 | 62.15 ± 2.85 | 86.10 ± 2.44 | 72.12 ± 1.22 |
|  | No condition events (I3) | 92.39 ± 0.15 | 74.25 ± 0.96 | 94.84 ± 0.27 | 98.25 ± 0.32 | 64.35 ± 0.58 | 80.56 ± 2.77 | 71.53 ± 1.03 |
|  | Generic surgery token (I4) | 94.20 ± 0.10 | 77.29 ± 0.46 | 94.99 ± 0.18 | 98.43 ± 0.24 | 64.30 ± 0.66 | 82.13 ± 2.13 | 72.11 ± 0.65 |
| Interaction | Procedure-as-baseline + generic surgery (A3+I4) | 94.19 ± 0.16 | 76.60 ± 1.48 | 94.80 ± 0.24 | 98.24 ± 0.29 | 64.12 ± 1.22 | 80.35 ± 2.48 | 71.30 ± 1.05 |
| Values are mean ± SD across five fold-specific models (%). Each episode contributes one surgery-index prediction (n = 23,538; coded PO-AKI prevalence, 10.1%). Threshold-dependent metrics use a probability threshold of 0.50.  †The no-baseline-loss variant did not train the surgery-index prediction head; these metrics are shown only to demonstrate its dependence on explicit supervision.  ‡In one fold, the surgery-index head predicted no positive cases; precision and F1 were therefore set to 0 for that fold. | | | | | | | | |

| **Supplementary Table S5. Complete postoperative dynamic performance of STREAM ablation variants** | | | | | | | | |
| --- | --- | --- | --- | --- | --- | --- | --- | --- |
| **Category** | **Ablation** | **AUROC** | **AUPRC** | **Accuracy** | **Specificity** | **Sensitivity** | **Precision** | **F1** |
| Full model | — | 94.82 ± 0.09 | 58.86 ± 0.87 | 98.30 ± 0.12 | 99.17 ± 0.12 | 63.86 ± 2.61 | 61.64 ± 2.29 | 61.72 ± 1.39 |
| Architecture | No time encoding (A1) | 95.32 ± 0.45 | 59.27 ± 0.85 | 98.27 ± 0.17 | 99.16 ± 0.22 | 64.08 ± 2.33 | 59.66 ± 3.50 | 60.26 ± 1.42 |
|  | Unidirectional attention (A2) | 94.94 ± 0.29 | 58.31 ± 0.71 | 98.27 ± 0.06 | 99.18 ± 0.07 | 59.63 ± 2.45 | 60.66 ± 1.46 | 59.50 ± 1.20 |
|  | Procedure token as baseline (A3) | 94.51 ± 0.29 | 55.95 ± 1.28 | 98.16 ± 0.33 | 99.04 ± 0.36 | 60.35 ± 1.78 | 59.62 ± 4.12 | 59.44 ± 2.40 |
| Training objective | No baseline loss (A4) | 94.48 ± 0.49 | 56.39 ± 2.80 | 97.53 ± 1.15 | 98.36 ± 1.26 | 61.06 ± 3.95 | 54.66 ± 10.81 | 56.38 ± 5.91 |
|  | No MEM loss (A5) | 94.82 ± 0.29 | 58.20 ± 1.25 | 98.30 ± 0.19 | 99.18 ± 0.25 | 62.82 ± 2.70 | 62.04 ± 2.92 | 61.64 ± 0.81 |
| Input modality | No medication events (I1) | 88.83 ± 0.34 | 23.10 ± 0.77 | 96.28 ± 0.40 | 98.19 ± 0.57 | 24.41 ± 4.77 | 29.41 ± 4.73 | 25.14 ± 1.75 |
|  | No laboratory events (I2) | 94.64 ± 0.37 | 56.77 ± 2.05 | 97.39 ± 0.67 | 98.16 ± 0.78 | 62.46 ± 4.26 | 52.49 ± 7.75 | 56.02 ± 2.81 |
|  | No condition events (I3) | 89.58 ± 0.85 | 46.61 ± 0.66 | 95.95 ± 2.13 | 96.60 ± 2.29 | 69.18 ± 3.09 | 36.53 ± 11.27 | 44.41 ± 9.10 |
|  | Generic surgery token (I4) | 94.41 ± 0.30 | 58.20 ± 0.95 | 98.35 ± 0.11 | 99.29 ± 0.18 | 60.41 ± 5.39 | 63.24 ± 3.50 | 60.79 ± 1.63 |
| Interaction | Procedure-as-baseline + generic surgery (A3+I4) | 94.72 ± 0.55 | 57.47 ± 1.47 | 98.20 ± 0.45 | 99.09 ± 0.52 | 61.72 ± 3.06 | 61.18 ± 5.20 | 60.58 ± 2.34 |
| Values are mean ± SD across five fold-specific models (%). Postoperative metrics were calculated separately within each valid 6-hour interval and macro-averaged across intervals, as defined in the main Methods. Threshold-dependent metrics use a probability threshold of 0.50. | | | | | | | | |

**Robustness to unresolved within-timestamp ordering**

Within-timestamp ordering perturbations produced relatively small changes in model output overall (**Supplementary Table S6**). Across 116,425 shuffle evaluations from 23,285 eligible windows, the median absolute logit change was 0.032, and the mean was 0.154. Sensitivity was negligible for two-event ties and increased for larger simultaneous-event groups, with median absolute logit changes of approximately 0.049–0.057 for groups containing three or more events. These findings support permutation averaging for event-arrival attribution when multiple events share the same recorded timestamp.

| **Supplementary Table S6. Sensitivity to unresolved within-timestamp event ordering.** | | | | |
| --- | --- | --- | --- | --- |
| **Tie-group size** | **Shuffle evaluations, n** | **Median \|Δlogit\|** | **Mean \|Δlogit\|** | **Fraction with \|Δlogit\|>0.05** |
| Overall | 116,425 | 0.032 | 0.154 | 0.467 |
| 2 | 14,155 | ≈0 | 0.056 | 0.189 |
| 3–4 | 32,305 | 0.049 | 0.160 | 0.498 |
| 5–8 | 33,080 | 0.055 | 0.161 | 0.510 |
| 9–16 | 25,500 | 0.057 | 0.178 | 0.511 |
| ≥17 | 11,385 | 0.052 | 0.183 | 0.503 |

**Event representation and model configuration**

**Supplementary Table S7** summarizes the event vocabulary structure used by STREAM. Detailed event construction, encoding, and normalization procedures are described in the main Methods. The complete STREAM token vocabulary is provided separately as **Supplementary Data 1**.

| **Supplementary Table S7. Summary of the STREAM event vocabulary.** | | |
| --- | --- | --- |
| **Event Domain** | **Token Format** | **Encoding/Normalization** |
| **Demographics** | AGE, SEX\|omop_concept_id, RACE\|omop_concept_id | Age standardized within training fold; sex/race represented by OMOP concepts |
| **Conditions** | CCS\|ccs_codes | ICD-9-CM/ICD-10-CM to CCS |
| **Medications** | RXCUI\|rxcui, MED_OTHERS | NDC/RxNorm to ingredient-level RxCUI; start/end boundary encoded separately |
| **Laboratory Tests** | LAB\|omop_concept_id, LAB_OTHERS | Unit harmonization within each laboratory test; numerical values normalized within training fold |
| **Index Surgery** | SNOMED\|snomed_code | Procedure mapped to SNOMED CT |

**Supplementary Table S8** summarizes the final STREAM architecture and training configuration. Training and checkpoint-selection procedures are described in the main Methods.

| **Supplementary Table S8. STREAM architecture and training configuration.** | |
| --- | --- |
| **Setting** | **Value** |
| Optimizer | AdamW |
| Batch size | 16 |
| Initial learning rate | 1e-4 |
| Weight decay | 1e-2 |
| Dropout | 0.2 |
| Hidden dimension | 128 |
| Transformer layers | 2 |
| Attention heads | 4 |
| FFN dimension | 256 |
| Attention scheme | Structured block attention; postoperative unidirectional (past-only) |
| LR scheduler | StepLR, step=5, gamma=0.7 |
| MEM mask ratio | 0.15 |
| Loss weights | online 1.0, baseline 0.3, MEM 0.01 |

**Supplementary Table S9** summarizes the XGBoost configuration used for the static nonlinear comparator.

| **Supplementary Table S9. XGBoost configuration.** | |
| --- | --- |
| **Setting** | **Value** |
| max_depth | 6 |
| eta (learning rate) | 0.1 |
| subsample | 0.8 |
| colsample_bytree | 0.8 |
| min_child_weight | 1.0 |
| lambda (L2 penalty) | 1.0 |
| alpha (L1), gamma | 0, 0 |
| scale_pos_weight | 16.54–16.78 |
| eval_metric | auroc, auprc, logloss |
| num_boost_round | 500 |
| early_stopping_rounds | 30 |
| seed | 42 |
| Classification threshold | 0.5 |

**Representative patient-level event-arrival trajectories**

**Supplementary Figure S1** shows representative STREAM trajectories for one episode with coded PO-AKI and one episode without coded PO-AKI, illustrating how event-specific probability updates accumulated over postoperative time.

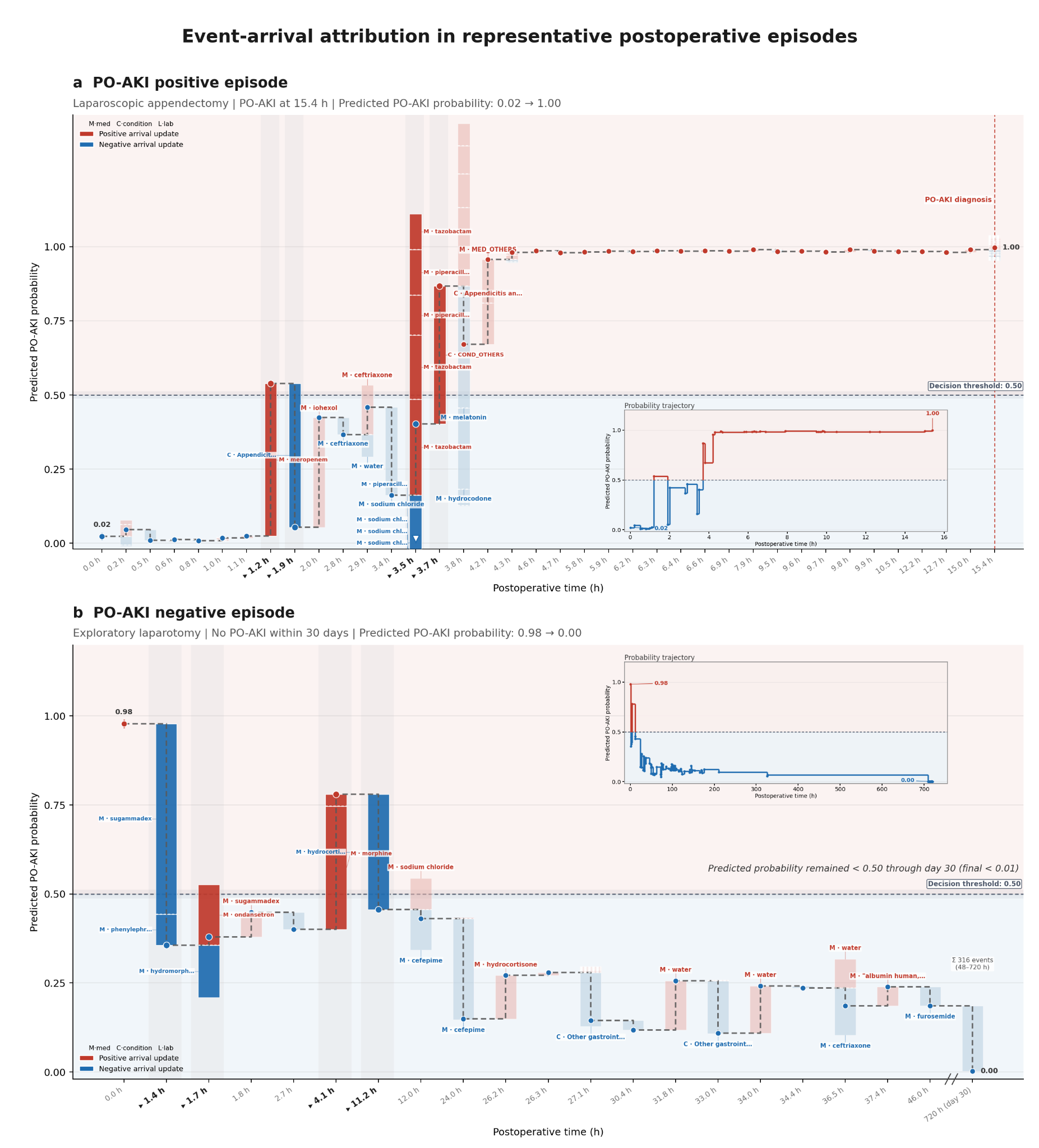

**Supplementary Figure S1. Patient-level event-arrival attribution for representative postoperative trajectories. (a)** PO-AKI-positive episode following laparoscopic appendectomy, with the first coded PO-AKI diagnosis at 15.4 hours after surgery. **(b)** PO-AKI-negative episode following exploratory laparotomy, with no coded PO-AKI within 30 days. Each red or blue bar spans the predicted probabilities before and after an individual event was incorporated into STREAM; red indicates an increase and blue a decrease in model-estimated PO-AKI risk. Darker shades mark updates that crossed the 0.50 threshold, whereas lighter shades mark updates that did not. The dashed trajectories show predicted probability after all events at each recorded timestamp were incorporated. Event contributions were accumulated on the logit scale before conversion to probability. For events sharing the same recorded timestamp, event-specific contributions were averaged across within-timestamp permutations. The horizontal dashed line at 0.50 is shown as a reference threshold for visualization. Insets summarize the complete postoperative risk trajectories, and the vertical dashed line in panel (**a)** marks the coded PO-AKI diagnosis time.

**Fixed-window contrast for representative patient trajectories**

**Supplementary Figure S2** presents matched Time-series Transformer trajectories for the same episodes, illustrating interval-level risk updates at predefined 6-hour boundaries.

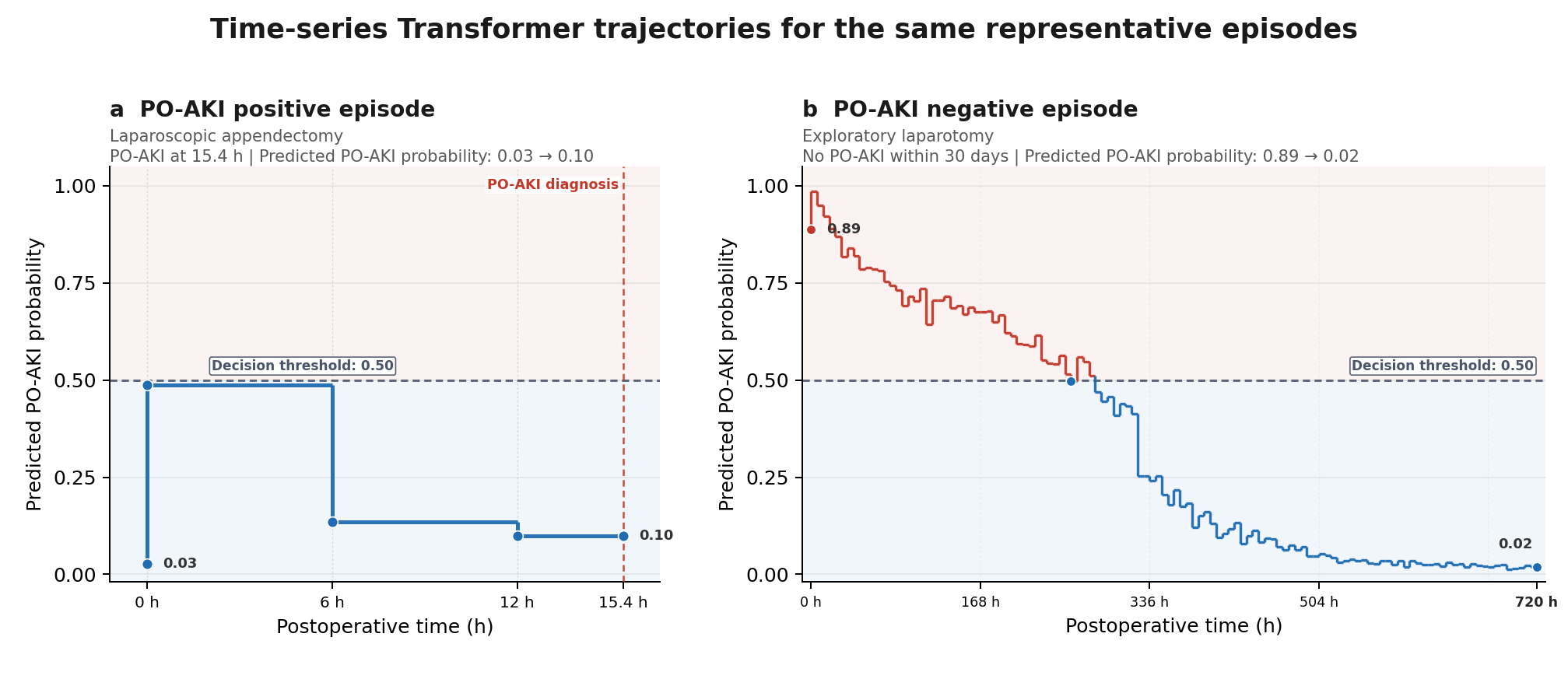

**Supplementary Figure S2. Time-series Transformer trajectories for the representative postoperative episodes shown in Supplementary Figure S1.** **(a)** Episode following laparoscopic appendectomy with the first coded PO-AKI diagnosis at 15.4 hours after surgery. **(b)** Episode following exploratory laparotomy with no coded PO-AKI within 30 days. Predicted PO-AKI probability was updated at fixed 6-hour intervals. The horizontal dashed line at 0.50 is shown as a visual reference, and the vertical dashed line in panel (**a)** marks the coded PO-AKI diagnosis time.

**Complementary model interpretation analyses**

Complementary analyses further characterize STREAM’s model behavior through temporal grouped deletion, age and procedure substitution, representative patient-level event-arrival trajectories, and matched fixed-window Time-series Transformer trajectories. Population-level event-arrival attribution and within-window event-order sensitivity are presented in the main manuscript.

**Supplementary Figure S3** summarizes three complementary perturbation analyses. Age substitution showed a monotonic increase in surgery-index PO-AKI logit with increasing age: relative to the reference age of 48 years, younger substituted ages produced lower logits, whereas older ages produced higher logits across the five fold-specific models, with the response flattening at older ages. Procedure substitution produced a broad range of surgery-index risk shifts across the 75 evaluated procedures and was positively associated with observed procedure-specific coded PO-AKI incidence (Spearman $\rho=0.667$).

Temporal grouped deletion showed that attribution patterns varied across the perioperative trajectory. Kidney and urinary tract disease, shock, diabetes with complications, and hypertension with complications showed positive attribution before surgery and generally retained positive attribution postoperatively, while respiratory failure or insufficiency emerged as a postoperative positive signal. Several diagnosis groups showed predominantly negative postoperative attribution, including acute myocardial infarction across multiple postoperative windows. Overall, attribution direction and magnitude could persist, emerge, or attenuate as the clinical trajectory evolved.

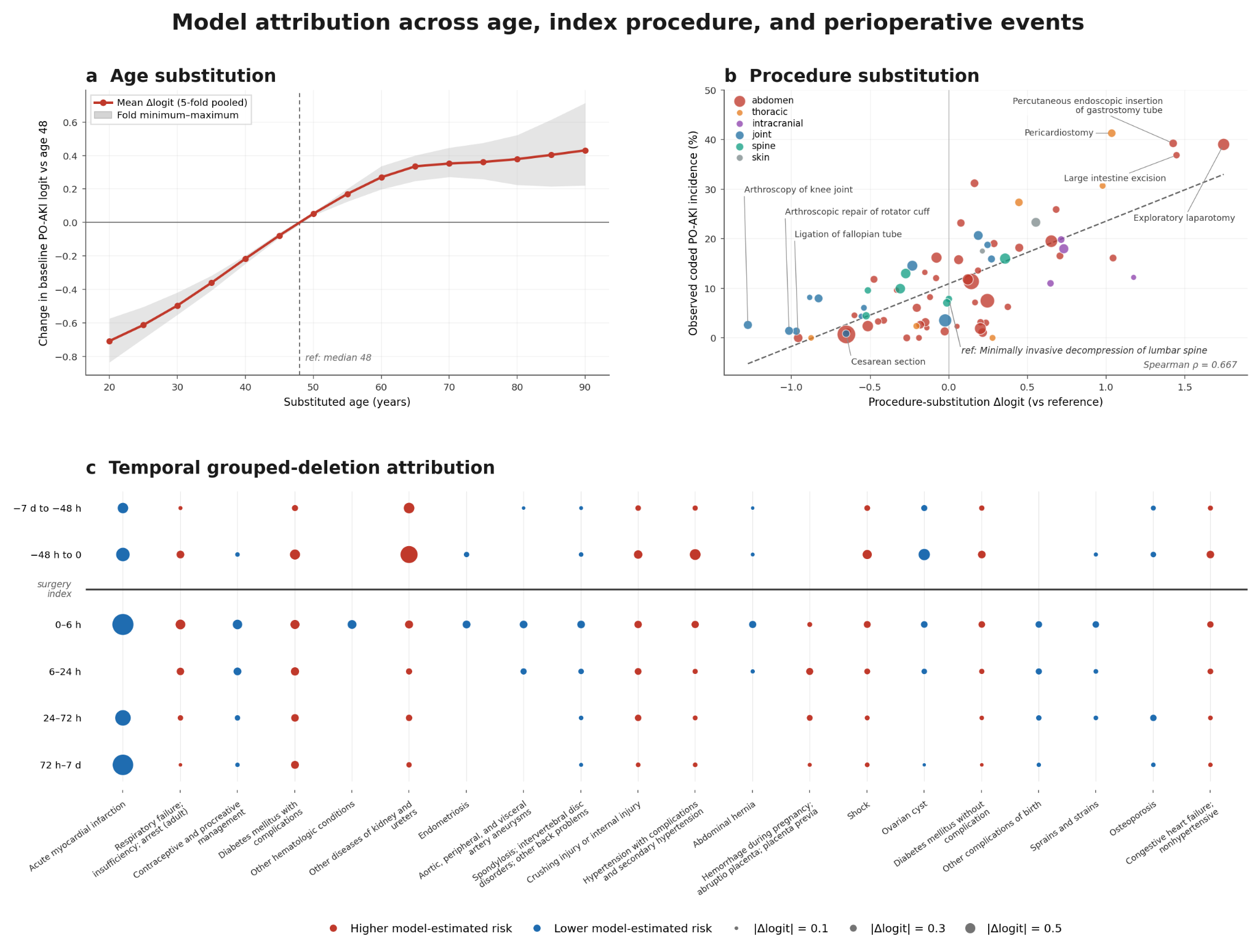

**Supplementary Figure S3. Model attribution across age, index procedure, and temporally grouped perioperative events.** **(a)** Within-episode age substitution showing the change in surgery-index PO-AKI logit relative to the reference age of 48 years. The line shows the five-fold mean, and the shaded band indicates the fold-wise minimum–maximum range. **(b)** Procedure-substitution analysis comparing procedure-specific change in surgery-index PO-AKI logit with observed procedure-specific coded PO-AKI incidence. Point size reflects procedure frequency, and color indicates surgical category; the dashed line summarizes the cross-procedure association (Spearman $\rho=0.667$). **(c)** Replicated temporal grouped-deletion attributions across perioperative time windows. Red and blue markers indicate higher and lower model-estimated risk attribution, respectively, and marker size represents $|\Delta logit|$.

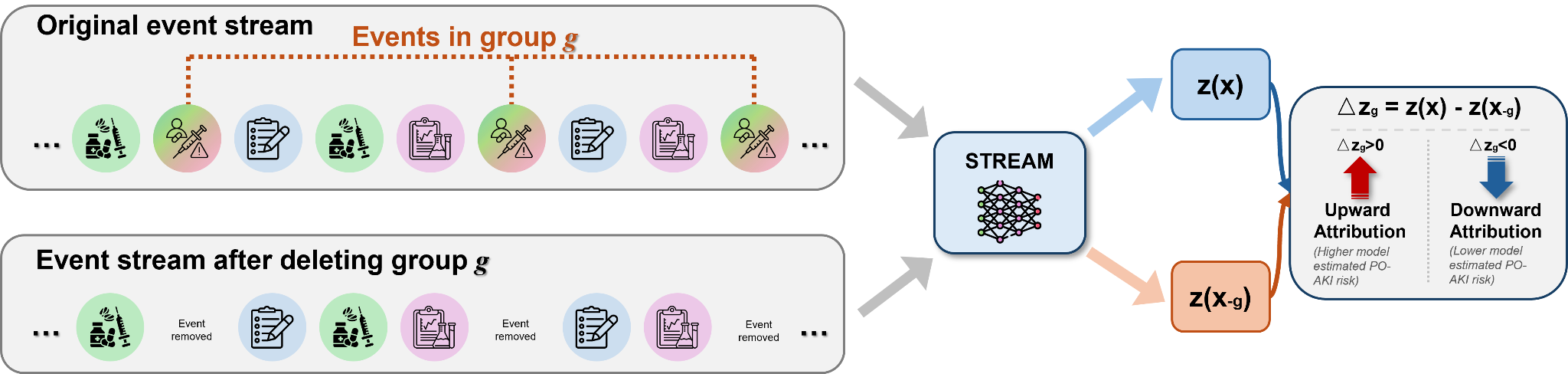

**Supplementary Figure S4. Temporal grouped-deletion attribution for STREAM.** Schematic of the grouped-deletion analysis described in the main Methods. Clinical events are grouped by modality, code, and perioperative time window, and attribution is summarized as the change in model-estimated PO-AKI logit after removal of the corresponding event group. Positive and negative values indicate upward and downward attribution, respectively.

**Supplementary Algorithm 1**

| ***Algorithm 1*** *Unified training and evaluation of fixed-window time-series baselines* | |
| --- | --- |
| 1 | **procedure TrainTimeSeriesBaseline(D_dev_, D_test_, model)** |
| 2 | Create five patient-grouped folds from D_dev_. |
| 3 | **for** fold = 1, …, 5 **do** |
| 4 | Assign one fold to validation and the remaining folds to training. |
| 5 | Fit preprocessing parameters, including normalization statistics and feature means μ, using training data only. |
| 6 | Construct training, validation, and test sequences X ∈ ℝ^(149×548): 28 pre-index positions, 1 surgery-index position, and 120 postoperative positions; retain M, Δ, and V. |
| 7 | Initialize a new model and separate surgery-index and postoperative prediction heads. |
| 8 | **for each** training epoch **do** |
| 9 | **for each** training mini-batch **do** |
| 10 | **if** model = GRU-D **then** |
| 11 | H ← GRU-D(X, M, Δ, μ). |
| 12 | **else** |
| 13 | X̃ ← Impute missing numerical values in X using causal LOCF, followed by μ when no earlier observation is available. |
| 14 | **if** model = LSTM **then** |
| 15 | H ← UnidirectionalLSTM(Project128(X̃)). |
| 16 | **else** [Time-series Transformer] |
| 17 | A ← BlockAttentionMask(pre ↔ pre; index ← pre + index; post ← index + current/prior post). |
| 18 | H ← Transformer2(Embed(X̃) + LearnedPositionEmbeddings, A). |
| 19 | **end if** |
| 20 | **end if** |
| 21 | p_BASE_ ← sigmoid(head_BASE_(H[28])). |
| 22 | p_k_ ← sigmoid(head_post_(H[28 + k])), k = 1, …, 120. |
| 23 | L ← λ_BASE_ · BCE(y, p_BASE_) + λ_post_ · MaskedBCE(y, {p_k_}, V). |
| 24 | Update model parameters by backpropagating L. |
| 25 | end for |
| 26 | Evaluate on validation data without parameter updates; retain the checkpoint maximizing 0.9 × AUPRC_online,macro_ + 0.1 × AUPRC_BASE_. |
| 27 | end for |
| 28 | Evaluate the selected checkpoint on the same held-out temporal test set D_test_. |
| 29 | end for |
| 30 | **return** mean (minimum, maximum) test metrics across the five fold-specific models. |
| 31 | end procedure |

*Notes:* Positions use zero-based indexing: 0–27 are pre-index positions, 28 is the surgery-index position, and 29–148 are postoperative positions. The surgery index is anchored at recorded surgery completion. M denotes observation masks; Δ denotes elapsed time since the last observation; V identifies valid postoperative prediction positions before the first coded PO-AKI diagnosis or through postoperative day 30 for episodes without coded PO-AKI. The label y indicates coded PO-AKI within 30 days. MaskedBCE averages postoperative binary cross-entropy over valid positions only; λ_BASE_ and λ_post_ are the prespecified loss weights. LOCF uses earlier observations only and is applied to numerical features. All training-derived preprocessing parameters are applied unchanged to validation and test data. AUPRC_online,macro_ is the unweighted mean AUPRC across valid 6-hour postoperative intervals.
